# Immune history influences SARS-CoV-2 booster impacts: the role of efficacy and redundancy

**DOI:** 10.1101/2024.03.06.24303879

**Authors:** Sophie L. Larsen, Iffat Noor, Haylee West, Eliana Chandra, Pamela P. Martinez, Alicia N. M. Kraay

## Abstract

Given the continued emergence of SARS-CoV-2 variants of concern as well as unprecedented vaccine development, it is crucial to understand the effect of the updated vaccine formulations at the population level. While bivalent formulations have higher efficacy in vaccine trials, translating these findings to real-world effectiveness is challenging due to the diversity in immune history, especially in settings with a high degree of natural immunity. Known socioeconomic disparities in key metrics such as vaccine coverage, social distancing, and access to healthcare have likely shaped the development and distribution of this immune landscape. Yet little has been done to investigate the impact of booster formulation in the context of host heterogeneity. Using two complementary mathematical models that capture host demographics and immune histories over time, we investigated the potential impacts of bivalent and monovalent boosters in low– and middle-income countries (LMICs). These models allowed us to test the role of natural immunity and cross-protection in determining the optimal booster strategy. Our results show that to avert deaths from a new variant in populations with high immune history, it is more important that a booster is implemented than which booster is implemented (bivalent vs. monovalent). However, in populations with low preexisting immunity, bivalent boosters can become optimal. These findings suggest that for many LMICs – where acquiring a new vaccine stock may be economically prohibitive – monovalent boosters can still be implemented as long as pre-existing immunity is high.

## Introduction

Host and pathogen heterogeneity are at the core of understanding infectious disease dynamics, including the potential benefit of intervention strategies like the original SARS-CoV-2 monovalent and updated bivalent booster vaccines. At the individual level, variation in behavior, viral shedding, and/or infectiousness can drive superspreading events across pathogens (e.g. [1, 2]). At the population level, host factors can also influence disease transmission. For instance, rent-to-income ratio and population density are associated with SARS-CoV-2 superspreading in Hong Kong, a so-called “double disadvantage” for impoverished individuals living in high-risk urban residential environments [3]. Lockdown mobility, testing, vaccination, and mortality during the SARS-CoV-2 pandemic have all been shown to be associated with socioeconomic status (SES) [4–6]. Beyond the host, some pathogens evolve over time, with broad impacts on transmission potential. Rapid evolution observed for SARS-CoV-2 resulted in the emergence of several variants distinct from the original wild-type virus, including Omicron. One early-pandemic estimate of the household secondary attack rate for SARS-CoV-2 was placed at 18.9%, but later rose to 42.7% for Omicron cases [7].

It is crucial to understand the interactions of these pathogen and host factors cumulatively, through time. As historical variants give way to their successors, experimental work with serological data has documented substantial differences in immune response to SARS-CoV-2 variants and vaccination by variant-specific immune history (e.g. [8–10]). This suggests the presence of an immune imprinting effect, where an individual’s prior exposure can impact the adaptive immune response to new infections [8, 11] – a phenomenon not only observed for SARS-CoV-2, but also in other respiratory viruses such as influenza and SARS-CoV-1 (e.g. [12, 13]). This signature is also present in mouse models when immunizing sequentially with SARS-CoV-2 and SARS-CoV-1 or endemic coronaviruses [14, 15]. Despite these serological findings, the impacts of SARS-CoV-2 imprinting at the population level are presently unknown. Given that host factors are known to have influenced transmission dynamics and protective behaviors throughout the pandemic ([3–5]), they also have the potential to shape variant history and the development of the immune landscape of the population. For example, an individual with low SES who was not able to social distance early in the pandemic [4] might be more likely to have had an early-pandemic infection prior to the emergence of new variants. On the other hand, a low SES individual is less likely to be vaccinated than a high SES counterpart [5]. Yet socioeconomic status is still a commonly unrecognized axis of host heterogeneity in disease modeling [16, 17].

Mathematical models can be wielded as a powerful tool for public health when host– and pathogen-level data are plentiful, but an understanding of complex population-level dynamics is lacking [18]. Understanding booster impacts under the influence of population and pathogen heterogeneity can inform not only the acquisition and implementation of booster vaccines but also shed light on future strategies for booster formulation. In this work, we synthesized serological measures of variant-specific immune history, socioeconomic disparities, temporal vaccination trends, and broad variation of historical variant wave sizes to inform models of transmission for three countries (India, Ecuador, and Malaysia) and evaluated possible landscapes of immunity more broadly across low– and middle-income country (LMIC) settings. Using this immune history, we forward-simulated the potential impacts of three formulations of bivalent or monovalent boosters in two models with complementary strengths, in the context of diverse landscapes of immunity and varying levels of pathogen immune escape or adaptation in infectiousness.

## Results

### Landscape of immunity

In order to characterize the effect of variant-specific immunity on population-level booster impact, we identified three discrete, historical waves of SARS-CoV-2 for our analysis – Wild-type (WT), Delta, and Omicron – using India, Ecuador, and Malaysia as benchmark countries due to the diverse sizes of their variant waves (Figure S1). We first implemented a model that tracks the variant-specific immune history of individuals at each time-point, captures relative wave size for each country (Figure S2, Table S1), and incorporates the observed temporal vaccination trends for each country and socioeconomic group (Figure S3, Table S2). We refer to this as the history-specific model or HSM, and under this model, an individual’s risks of infection are explicitly influenced by their specific immune history and the currently circulating variant, while the infection fatality rate is influenced by immune history, socioeconomic status, and age (Figure 1A, Table S3).

**Figure 1:**
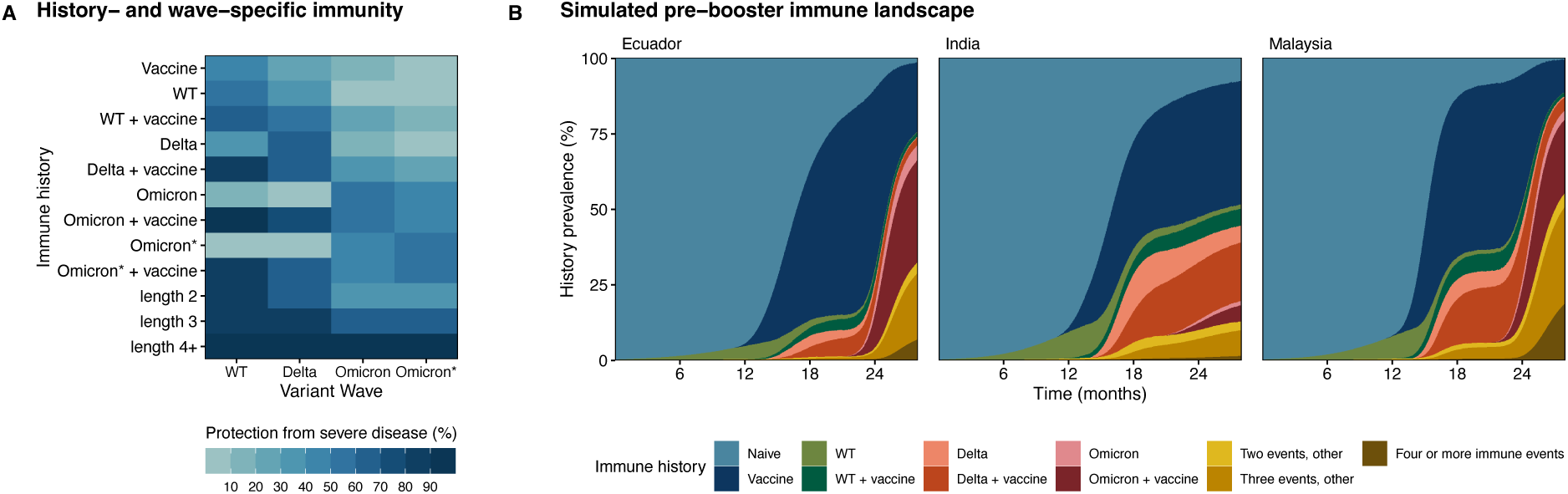
Landscapes of immunity from the history-specific model (HSM). (A) Level of protection from severe disease by immune history and variant wave, estimated using neutralizing antibody titers in human sera [19, 20]. Lengths 2, 3, and 4+ represent permutations of immune history that were not explicitly tracked (e.g. ‘WT + Delta + Omicron’). (B) Average immune history trajectories of 500 runs informed by historical incidence data from Ecuador, India, and Malaysia shown in Figure S1 [21]. Simulations of the prevalence of histories are shown until day 839, one day before booster vaccinations are implemented at 28 months. The infections over time for each country are shown in Figure S2. General simulation parameters are shown in Table S3, with country-specific population structure in Table S4, contact rates by country and SES in Table S5, wave– and country-specific stringency in Table S1, and vaccination parameters by country and SES in Table S2. The HSM compartmental diagram is shown in Figure S5.

Using the variant-specific immune histories of individuals tracked through historical simulations in the HSM, we explored the possible immune landscapes over time for the three countries prior to a booster intervention (Figure 1B) and inferred the prevalence of each history type prior to the time of boosting at 28 months (Figure S4). Our findings suggest that India had the highest share of naive individuals, with an average of 7.8% of the population having never been infected or vaccinated. Their vaccine-only population was also high (40.9%) compared to Ecuador (23.1%) and Malaysia (10.9%). Yet, despite having lower natural exposure overall, our simulations indicate that India carried by far the highest percentage of ‘Delta’ and ‘Delta + vaccine’ histories, at values of 5.6% and 19.4% respectively. In Malaysia, ‘Delta’ exposures appeared to be less common, but there was a much higher presence of ‘Omicron’ histories than in India (3.0% for ‘Omicron’ and 24.3% for ‘Omicron + vaccine’). A pattern similar to that of Malaysia was present in Ecuador, where Omicron histories were also more prevalent than Delta or WT. We used these diverse immune histories, particularly in comparing vaccine-only versus hybrid immunity, to assess the population-level of protection from a booster intervention under varying hypotheses of individual-level efficacy.

### Impact of booster formulation on disease incidence and deaths

We introduced booster vaccines at 28 months under three formulations – bivalent (WT + Omicron), monovalent (WT), and a hypothetical monovalent (Omicron) – and considered two possible scenarios of pathogen infectiousness (Table S3): one that is conservative (110% relative to Omicron) and another that more closely resembles previous increases in the observed secondary attack rate (SAR) between variants (130% relative to Omicron), considering that the SAR for Delta was *~* 60% higher than early-pandemic estimates and the SAR for Omicron was *~* 40% higher than for Delta [7]. To model protection conferred by these boosters, we considered a ‘history-dependent’ scenario where the impact of each formulation is based on an individual’s prior exposure, and calculated relative to history-specific responses to primary series vaccination (Figure S6). At 30 months, we introduced the Omicron* variant and updated history-specific immunity for both boosted and unboosted individuals (Figures 1A, 2A). Unsurprisingly, our results show that boosting always reduced cases (Figure 2B) and disease-related deaths (Figure 2C) in all three countries during this new wave, regardless of formulation. For example, in the 130% infectiousness scenario, wave peaks with boosting were 22-37% lower than the peak with no boosting in India, 33-39% in Ecuador, and 44-45% in Malaysia. The smaller reduction observed in India may be attributable to a lower boosting rate (Figure S3) and the distinct immune landscape compared to Malaysia and Ecuador (Figure 1B).

**Figure 2:**
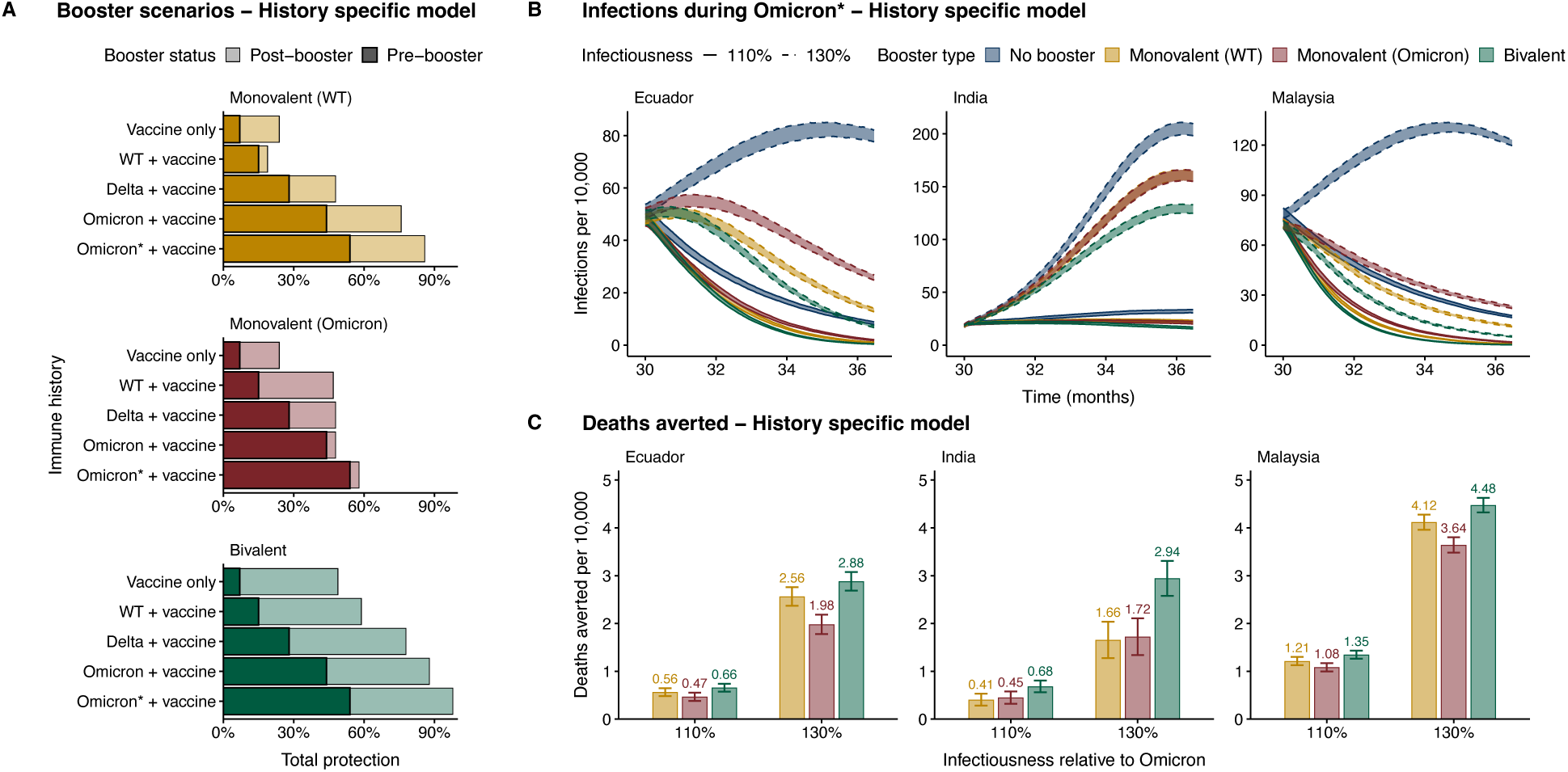
HSM booster parameters and projections during the Omicron* period. (A) Booster protection against severe disease during the Omicron* wave, by immune history, under a bivalent, WT monovalent, or hypothetical Omicron monovalent formulation. (B) Infection trends under three boosters or a no-boosting scenario based on the characteristics of Ecuador, India, and Malaysia. We simulated two scenarios of infectiousness where Omicron* is 10% or 30% more infectious than Omicron. 95% confidence intervals from the t-distribution are shown (ribbons). (C) Deaths averted by boosting since the start of Omicron* (30 months) through the end of simulations, under each booster. 95% confidence intervals from the t-distribution are shown with whiskers. Boosting parameters, which are assumed to be 10 weeks faster than for primary series vaccination, are shown in Table S8.

In order to quantify the difference in deaths averted during the Omicron* wave across booster formulations (Figure 2C), we estimated the relative benefit of switching from monovalent (WT) to monovalent (Omicron) or to bivalent (Equation 1) specific to each country and scenario. Values *<* 1 represent a net loss in deaths averted from changing booster formulations, whereas values *>* 1 represent that the new formulation averts more deaths than the previous formulation. If the benefit of switching from monovalent (WT) to a different formulation is greater than the original benefit of implementing the monovalent (WT), then the relative benefit will be greater than 2. Therefore, if the relative benefit is in the interval (1, 2), then the new formulation is still optimal to the monovalent (WT) booster in the given scenario, but the effect of boosting on the deaths averted is stronger than the effect of booster formulation.

When comparing the relative benefit of switching to monovalent (Omicron), India was the only country that would slightly benefit from this hypothetical booster, represented by values greater than 1.0 in both the 110% and 130% more infectious scenarios (Table S6). This marginal benefit in India may be attributable to the high prevalence of Omicron histories in Ecuador and Malaysia compared to India, where Delta histories appeared to be more prevalent (Figure 1B). When looking at the case of bivalent boosters, we found that in the 110% infectiousness scenario, the relative benefit of bivalent boosters was 1.12, 1.18, and 1.66, for Malaysia, Ecuador, and India, respectively. For the 130% infectiousness scenario, the relative benefit of bivalent boosters was 1.09 for Malaysia, 1.12 for Ecuador, and 1.77 for India (Table S6).

This surprising finding – that switching formulations from monovalent (WT) to bivalent in a given scenario always yielded diminishing returns on the deaths averted with relative benefit values less than 2.0 – was consistent even in scenarios with conservative booster rollout speed (Figures S3E, S7, Table S7), and in scenarios where booster curves for all countries match the superior rollout trajectory seen in Malaysia (Figure S8, Table S7). In order to isolate the effects of the boosters, the main results show findings when primary-series vaccination is stopped at the end of the Omicron wave. When we relaxed this assumption by continuing primary series vaccination through Omicron*, we found that this did not cause the relative benefit of the bivalent to surpass 2.0 (Figures S3C, S9, Table S7). Finally, while our history-specific booster scenarios were informed by responses to primary series vaccination, they were hypothetical. We therefore tested two additional scenarios for booster efficacy: (1) same efficacy, where all individuals received the same numeric change in protection from a given booster (Figure S10), and (2) same endpoint, where all individuals reached the same end level of protection from boosting (Figure S11). The conclusion about the relative benefit of bivalent boosters remained consistent (Table S7).

### Beyond country-specific trajectories

In order to explore a wider range of scenarios that capture the relative influence of preexisting immunity and degree of overlap with the currently circulating variant, we implemented a complementary and flexible model, which we call the hybrid-immunity model (HIM). The HIM is a compartmental model that tracks prior infection (none, one or more; not variant-specific), vaccine history (none, primary series, primary series+booster), and simulates expected cases and deaths during the Omicron* wave. For the three bench-marking countries, the level of cross-protection, prior infection, and baseline transmission rates expected are parameterized based on the outputs of the history-specific model prior to boosting (Figures 1B, S4, Table S1, S9). Country-specific booster trends are also matched (Figure S12). This allowed us to compare the predictions between the two models for the benchmarked countries. We then used the hybrid immunity model to vary these dimensions of immunity and to more thoroughly explore how booster impacts might be influenced by prior population infection and the degree of cross protection conferred by natural infection. To determine prior infections and cross protection at the start of the HIM simulations on day 900, we used immune history distributions from day 899 in the HSM under a no-boosting scenario (Figure S13); these may overestimate the cross protection, but because there is limited booster impact on the number of infections during the window from day 839 to 899 (Figure S2), this is a close approximation.

In general, projections of infections during Omicron* for the three countries were qualitatively similar for the HIM compared with the HSM, with similar outbreaks (Figure 3A). For the 110% infectiousness scenario, the HIM predicted small outbreaks in the three countries. All three countries experienced outbreaks in the 130% infectiousness scenario, with the biggest peaks in Malaysia and India. Boosting reduced the size of the Omicron* wave across all three countries, with bivalent boosters having a stronger impact than monovalent boosters. However, similar to the HSM, the impact of vaccine formulation was small. When looking at deaths across the three countries, models revealed substantial differences across countries in the potential for booster impacts, with India having by far the highest number of deaths with or without boosting, matching the HSM (Figures S14, S15). This split in deaths across countries was larger overall in the HIM than in the HSM, and additionally, the HIM predicted smaller outbreaks for Malaysia than the HSM. As a result India was predicted to have the strongest potential benefit of boosting on deaths averted per 10,000 (Figures 3B (black dots), S16, Table S11). Like the HSM, these findings from the HIM were similar in the conservative booster rollout case (Figures S16, S17), but overall deaths were higher with slower booster rollout (Figure S14). We also tested scenarios where boosting started at the beginning of the Omicron* wave (Figure S18), with similar findings but lower deaths averted overall (Figures S19, S20).

**Figure 3:**
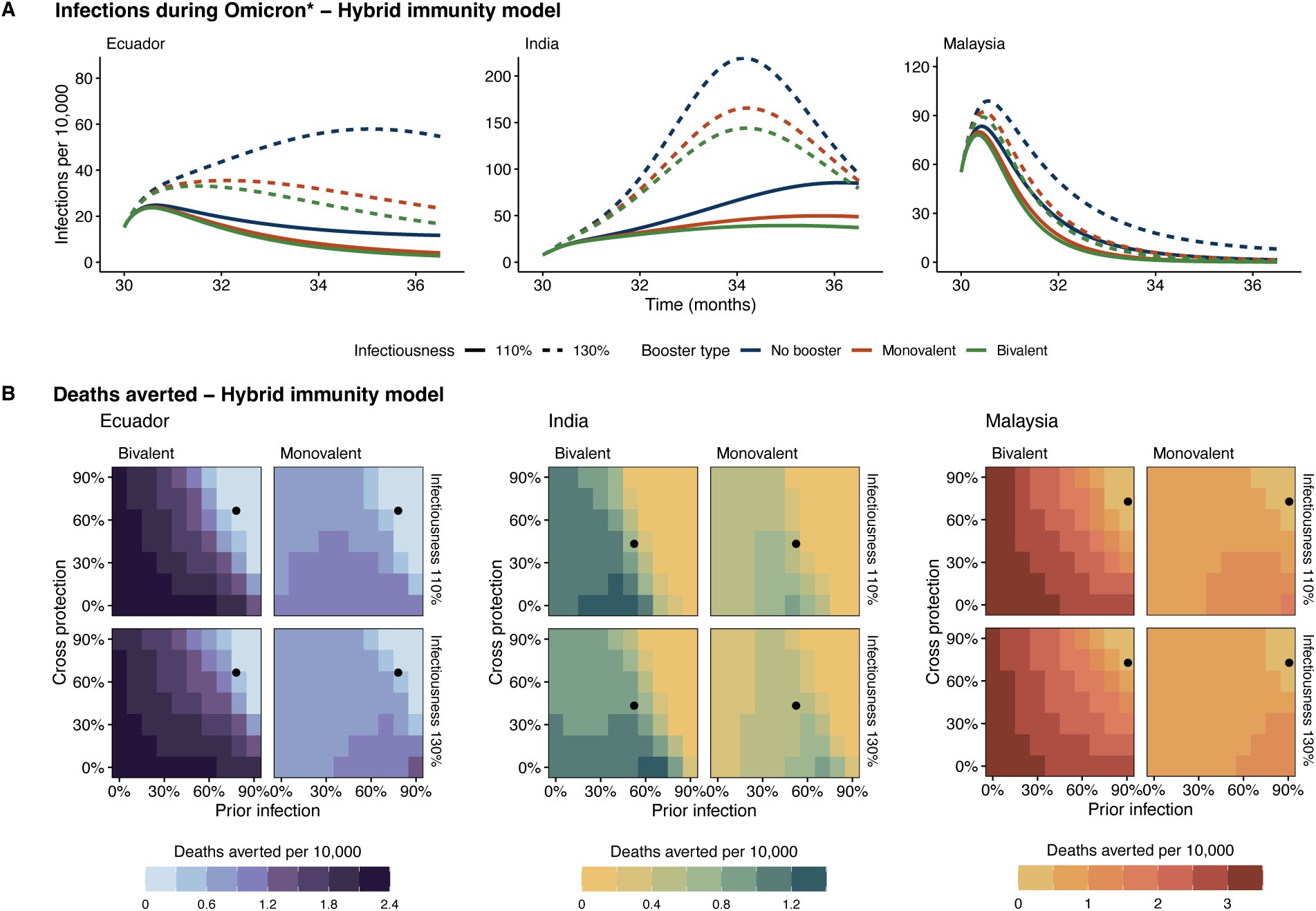
HIM projections. (A) Infections per 10,000 during the Omicron* period, where Omicron* is considered to be 10% or 30% more infectious than Omicron. (B) Deaths averted per 10,000 under a bivalent or monovalent booster, compared to a ‘no boosting’ scenario. Prior immunity represents the percentage of the population that has been previously infected. Cross protection represents the overlap between the population’s immune history and the currently circulating variant. Country-specific immunity levels are included (black dots). The HIM model structure is shown in Figure S5. General parameters are shown in Table S3, stringency in Table S1, initial conditions in Table S10, cross-protection in Table S9, and boosting parameters in Table S8.

### Consequences of immune escape and cross protection in diverse immunity contexts

Broader sweeps across the immune landscape using the HIM revealed that the similarity between monovalent and bivalent booster performance was largely determined by population immunity (Figure 3B, Figure 4). Generally, the gains from implementing a bivalent booster were most pronounced at low levels of population immunity, but absolute differences remained relatively small for all parameter values considered. The difference between monovalent and bivalent vaccination was smallest at high levels of cross protection and prior exposure (as was seen in the three benchmarking countries), and diverged more as the protection from natural infection decreased across both dimensions. For example, in Malaysia, the HSM estimated that 90.7% of the population had been infected by the start of Omicron* and that this protection conferred a 72.8% protection against disease-related death (Table S9). At that preexisting immunity level in a 130% infectiousness scenario, monovalent boosting was expected to avert 0.14 deaths per 10,000 compared with 0.22 deaths per 10,000 in the bivalent vaccination scenario, representing that the additional gain in deaths averted from jumping to an improved formulation (0.08) is just over half of what was gained when moving from no-boosting to monovalent – a relative benefit of 1.57 (Equation 1, Figure 4). These diminishing returns account for much of the high-immunity parameter space. However, in the absence of any baseline immunity, bivalent boosting could avert 3.4 deaths per 10,000 compared with 0.6 deaths per 10,000 in the monovalent case. This gain of 2.8 deaths averted per 10,000 by improving formulations, compared to 0.6 gained by simply boosting, represents a relative benefit of 5.67. In the other countries, diminishing returns accounted for a wider swath of the parameter space (Figure 4). In general, at hypothetical lower levels of immunity, Malaysia was predicted to have higher impacts than the other countries (Figure 3B), reflecting that booster curves for Malaysia were superior (Figure S12). Whereas the total deaths averted were similarly sensitive to both the cross protection and prior infections (Figure 3B), the prevalence of prior infection was the greatest determining factor in whether a bivalent booster would be optimal.

**Figure 4:**
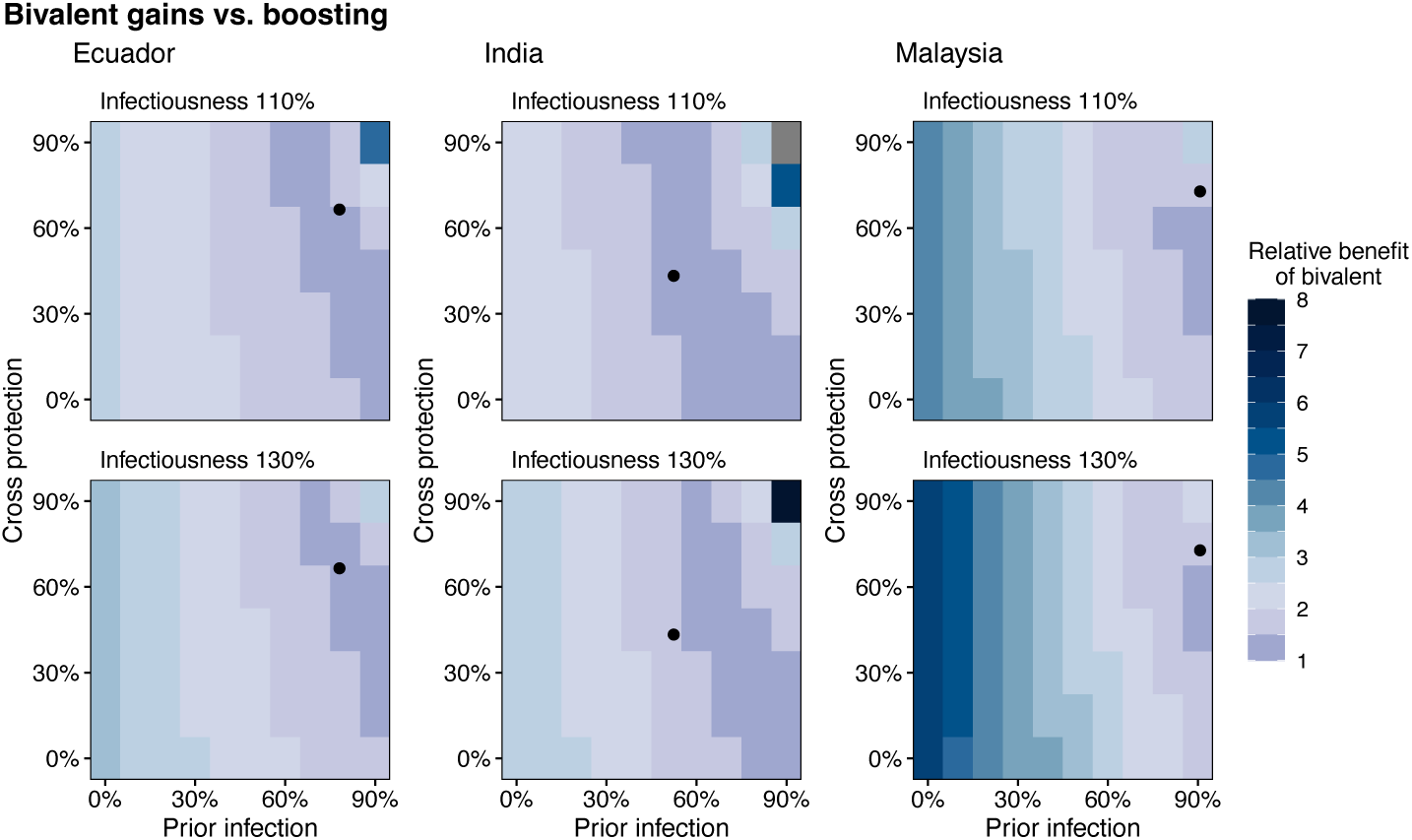
Comparison of bivalent vs. monovalent boosters in the HIM. Relative benefit of the bivalent booster is calculated with Equation 1, matching the HSM. Grey values represent high-immunity areas where zero deaths were averted by a monovalent booster and thus Equation 1 cannot be calculated. Prior immunity represents the percentage of the population that has been previously infected. Cross protection represents the overlap between the population’s immune history and the currently circulating variant. Country-specific immunity levels are included (black dots).

The HSM and HIM both assumed that individuals with pre-existing immunity have strongly reduced infectiousness based on [22], but this may not always be the case, particularly as new variants emerge. We thus tested an additional scenario in the hybrid immunity model where prior exposure or immunization had no impact on host infectiousness during the Omicron* wave, under two parameterizations. First, we left all other parameters related to the force of infection unchanged, including the baseline risk of infection given contact, but removed all transmission-reducing effects during Omicron* so that the effective infectious proportion is equal to the total infectious proportion. In this extreme scenario, all three countries experienced large waves regardless of whether the baseline infectiousness for Omicron* was 10% or 30% greater than Omicron (Figure S21). Under these conditions, it was almost always optimal to implement a bivalent booster (Figure S22). However, stringency in the HSM and HIM was calibrated to observed wave sizes for Omicron *with* the assumption of transmission-reducing immunity, so this scenario represents an extreme jump in the overall force of infection. We therefore ran an additional analysis where the overall force of infection was re-calibrated to fit the *R*_0_ of Omicron (Table S3) without transmission-reducing effects (Figures S23, S24). In this scenario, cases were still increased but the relative benefit of bivalent boosting was less than 2.0 in most of the high-immunity parameter space, and in all but one country-specific scenario (India, 130% more infectious). In general, HIM-projected deaths tended to be lower than the HSM across sensitivity analyses (Figures S14, S25, S26) – except in the case where transmission structure was adjusted but β was not recalibrated (Figure S27) – reflecting differences in the structure of immunity; the HSM is designed to capture more complex dynamics of immunity over time, number, and type of exposures, whereas the HIM tracks recent recovery (sterilizing immunity) and tiers of susceptibility depending on vaccination status and infection-naive versus previously infected. This was also reflected in the scale of the deaths averted which were generally lower across scenarios (Table 1).

**Table 1:** Comparison of HIM deaths per 10,000 across three scenarios: (1) Matching the HSM (column 4; as shown in Figures 3 and 4) (2) Removing the reduction in transmission for people with pre-existing immunity (column 5), and (3) Removing the transmission reduction but recalibrating β to remain consistent with Omicron [23] (column 6).

| Country | SAR | Booster | Match HSM | Adjust Transmission | Transmission Recalibrated |
| --- | --- | --- | --- | --- | --- |
| Ecuador | 110% | Monovalent | 0.07 | 0.3 | 0.57 |
|  | 110% | Bivalent | 0.11 | 0.94 | 0.84 |
|  | 130% | Monovalent | 0.27 | 0.24 | 0.59 |
|  | 130% | Bivalent | 0.38 | 0.88 | 1.02 |
| India | 110% | Monovalent | 0.4 | 0.13 | 0.67 |
|  | 110% | Bivalent | 0.57 | 0.61 | 1.14 |
|  | 130% | Monovalent | 0.58 | 0.1 | 0.43 |
|  | 130% | Bivalent | 0.96 | 0.58 | 0.9 |
| Malaysia | 110% | Monovalent | 0.06 | 0.32 | 0.45 |
|  | 110% | Bivalent | 0.11 | 1 | 0.6 |
|  | 130% | Monovalent | 0.14 | 0.28 | 0.57 |
|  | 130% | Bivalent | 0.22 | 1 | 0.86 |

Finally, in the HIM, we assume that 50% of individuals with prior infection start in the recovered (R) compartment due to the short interval between transmission waves. The remainder start in the susceptible but previously infected compartment. This assumption is roughly consistent with most of current population immunity having been gained during the Omicron wave (like in Malaysia and Ecuador), of which roughly half would have waned at the start of the Omicron* wave. Thus, at very high levels of prior immunity, a substantial fraction of the population cannot be reinfected until waning occurs, which could influence our estimates of booster impact. Additionally, this assumption may be less realistic for India, which experienced a larger Delta wave than Omicron (Figures S1, S2). As a sensitivity analysis, we re-ran our model assuming that only 25% of individuals with prior infection start in the R compartment. While cases and deaths were higher (Figures S28A, S29), the overall findings were unchanged (Figures S28, S30).

## Discussion

The continued emergence of new variants of SARS-CoV-2 poses a challenge for maintaining effective vaccine-induced immunity in the population. As individuals continue to acquire diverse immune histories that confer highly varied protection against newer variants [8], understanding the impact of old and new vaccine formulations in populations with varying exposure histories is crucial. We found that at the population level, particularly in the current context where population immunity is high, it is more important that a booster is implemented, than whether that booster is bivalent or monovalent. Strikingly, this was largely consistent across all three countries in both models, even in scenarios where rollout speed was conservative or an emerging variant was highly transmissible. While bivalent boosters showed a greater benefit when the rate of previous SARS-CoV-2 infections was low, those scenarios are unlikely for most countries. For example, a meta-analysis found that the rate of infection-induced seroprevalence in India may have been higher than 50% by the third quarter of 2021 [24], a year before bivalent boosters became available. In Malaysia, estimates from reported cases and age-stratified case fatality rates suggest that approximately 33% of individuals had been infected by December 2021, with only 23% of cases estimated to be reported [25]. Infection-induced seroprevalence estimates for a selection of European low– and middle-income countries (LMICs) had surpassed 50% by as early as November 2020 [24]. The continued benefit of the original monovalent boosters in these contexts is an encouraging finding, especially given that it may not be economically viable for an LMIC to acquire an entirely new vaccine stock. Still, care should be taken in interpreting the effect size of formulation across countries with varying population sizes, since 1 death averted per 10,000 is a larger absolute number in India than in Ecuador. Policymakers should consider the balance between relative and absolute gains when deciding which formulation to implement.

Monovalent boosters targeting the most recently circulating variant did not perform better than WT monovalent formulations in populations with a high degree of natural exposure. Red Queen dynamics [26] as well as documented imprinting effects [8] may give rise to an endless race to capture the newest variant match. While exploring boosters that directly target emerging variants is beyond the scope of this study, this has been the case for influenza virus. Influenza vaccine development and strain selection are based on what might be circulating during the upcoming season – informed by predictive models [27, 28] – and there can be severe impacts on vaccine efficacy if expectations do not align with reality [27]. In the face of this, a possible future direction for SARS-CoV-2 to make population-level gains in immunity is the development of pan-sarbecovirus or pan-coronavirus vaccines that induce broad cross-reactivity [29, 30]. Indeed, experimental work is already underway that supports this goal, but with a potentially long road ahead [29, 30].

Several additional factors should be considered when interpreting the findings of this paper. First, this work was conducted in response to a call from the World Health Organization in August 2022, and therefore our models and assumptions were built between 2022 and 2023. During a new variant wave, assumptions on the force of infection, rate of booster uptake, level of immune escape, and other factors are exploratory and hypothetical. We acknowledge that our simulations of Omicron* may be different from currently circulating variants and newly available boosters, but exploring post-Omicron lineages is beyond the scope of this study. Second, while neutralizing antibody titers allowed us to parameterize variant-specific protection in a way that could not be done with existing clinical trial data, and correlation with protection has been previously suggested for SARS-CoV-2 [31], we acknowledge that titers may not always be a good correlate of protection in all cases (e.g. [32]). Conversely, in the hybrid-immunity model, the impacts of a booster might be underestimated by using generalized booster efficacy and not accounting for the specific type of hybrid immunity. Third, while we chose three countries with distinct parameterizations to calibrate these models, fitting our models to data is challenging due to the difficulties of fitting individual-based models and incorporating individual immune trajectories to compartmental models. While our results are therefore more qualitative, they still match the patterns observed in incidence data.

When comparing the two models, the hybrid-immunity model might produce lower estimates of incidence and deaths than the history-specific model for several reasons – for example, the inclusion of age-specific contact rates (which are lower for older adults), reduced specificity of immune histories, and because immunity acquired during the current wave is likely to be stronger than pre-existing immunity, causing the cross-protection achieved by natural infection to change gradually over time during the Omicron* wave for the HSM but is fixed for the HIM. Deaths averted predicted by the HIM might also be lower than the HSM because booster vaccination does not confer even short term sterilizing immunity and only reduces probability of infection and severe disease for future exposures. While we acknowledge the limitations of our study, the outcomes of the two models presented here are consistent when evaluating the relative benefit of booster formulation and we are confident that the qualitative findings of relative booster impacts are robust.

In summary, incorporating numerous sources of host heterogeneity across diverse settings and alternative model structures has enabled us to quantify the relative impact of boosting compared to booster formulation. We have demonstrated a consistent finding that in some places, the original generation of SARS-CoV-2 monovalent boosters can still avert a similar number of deaths to the bivalent, without requiring the acquisition of a new vaccine stock. These results have potential implications for future vaccine policy and development.

## Methods

### History-specific model (HSM)

#### Immune histories

We projected previously reported data – neutralizing antibody titers from human sera which were stratified by immune history [19, 20] – onto a scale from 0.05-0.95, with larger numbers representing higher protection against SARS-CoV-2 (Figure 1A). Unvaccinated titers were those from individuals with no prior vaccine doses, and vaccinated titers were those from individuals with 2 or more doses (fully vaccinated).

The base probabilities of (1) being infected after contact and (2) dying due to infections are scaled down based on these wave– and history-specific parameters. This allowed us to parameterize single-infection histories as well as hybrid histories for the WT, Delta, and Omicron waves. For individuals with 2 prior infections, which was not accounted for in this data, we took an average of the ‘strain + vaccine’ serotypes for each wave. For four or more immune history events, we assumed a protection parameter of 0.05 (95%). For three history events (three infections or two infections + vaccine), we assumed the protection would fall halfway between 2 events and 4+ events for each wave. Finally, we constructed an Omicron* wave by drifting protection down by 0.10 for all groups, equal to the change in protection moving from the WT wave to Delta. Individuals with ‘Omicron*’ or ‘Omicron* + vaccine’ histories are given protection equal to what ‘Omicron’ and ‘Omicron + vaccine’ have during the Omicron wave.

#### Model structure

We implemented a stochastic, individual-based transmission model in Python. This model spans a period of three years, through 4 discrete waves of SARS-CoV-2 variants: WT (beginning in March 2020), Delta, Omicron, and a hypothetical Omicron lineage called Omicron* which we modeled to begin transmitting in October 2022. Each wave has a distinct risk of infection given contact, estimated from household secondary attack rates [7]. The simulated populations are grouped into children (0-20), adults (21-65), and elderly (older than 65) whose proportions reflect the age structure of each country [33], with 50% being high SES and 50% low SES. To capture relative waves sizes for each country, we implemented a stringency index with a value for each wave (Table S1). We ran 500 replicates for each set of simulation parameters. Simulations progress though a modified tau-leap algorithm [34]. General parameters are shown in Tables S3.

Country-specific, SES-stratified contact rates in this model are estimated from pre-pandemic contact data for each country [35] combined with estimates of unequal mobility by SES during the pandemic [4] and the tendency towards within-SES contact vs. across-SES [36, 37] (Table S5). Susceptible (*S*) individuals can become exposed (*E*) through contact with an infectious individual (*I*). The probability of becoming exposed given contact is stratified by variant-specific immune history. After exposure, they become infectious. The probability that an infected individual will recover or die is stratified by age and SES [4], as well as variant-specific immune history. While recently recovered (*R*), an individual does not have sterilizing immunity, but rather a peak level of protection dictated by their immune history (Figure 1A), which comes from neutralizing antibody titers in human sera [19, 20], and is order-agnostic. Recently recovered individuals have a lower risk of infection and disease-related death than those in the susceptible class. Here, the protection against infection is assumed to be 80% of the values shown in (Figure 1A) which apply to the risk of death – reflecting evidence that in the early months after SARS-CoV-2 infection or vaccination, protection against both severe disease and infection is high, but protection against infection is moderately lower [38]. After recovery, individuals eventually wane into the susceptible class again, with a dampened immune history protection parameter, and the amount of this dampening is dictated by their previous number of immune exposures – with more exposure resulting in a lower degree of waning (Table S3). The incorporation of peak (recently recovered class) and waned (susceptible class) protection as opposed to sterilizing immunity and susceptibility reflects evidence that individuals are not fully protected against reinfection even when their immune history is fresh [38]. This granularity in considering the peak and waned protection of an individual, across immune histories, is a core strength of this model.

#### Vaccination

SES-stratified vaccination is implemented from day 320 (during the WT wave, around January 2021) until the beginning of Omicron*, according to the true rates observed for each country [5] (Figure S3, Table S2). Susceptible, exposed, and recently recovered individuals can be vaccinated – and if they are not already recently recovered, they move to the recently recovered class. We stopped primary series vaccination at the start of Omicron* reflecting that a small number of primary series doses were projected to be given during Omicron* (Fig. S3), and to isolate the effect of boosting from interference by the primary series vaccines.

#### Boosting

Boosters function similarly to vaccination, but with faster timing and lower coverage (Figure S3, Table S8). Booster rollout was assumed to be 10 weeks faster than primary series vaccination, to reflect possible improvements in infrastructure (Figure S3D, Table S8). We assumed that the coverage of monovalent boosters in October 2023 represented a peak booster coverage level in the population [39]. Children and unvaccinated individuals are not eligible to receive a booster, reflecting booster eligibility at the time of the study.

Data on booster efficacy by variant-specific immune history and formulation are limited. We therefore used the neutralizing titer data [19, 20] to project booster-related changes in the level of protection against severe disease, using responses to primary-series vaccination during the WT wave. Because Omicron* is a hypothetical variant with no data on primary series vaccination responses, we assumed that the change in protection from boosting would be the same as for the Omicron variant. Bivalent boosters were assumed to confer the same numeric change as monovalent vaccines did during the WT wave. We considered monovalent boosters to have 40% of the efficacy of bivalent boosters against Omicron* infection based on estimates of efficacy against severe infection [40]. Because we started boosting two months prior to Omicron*, there was a two-month period when the bivalent and Omicron monovalent formulations were matched to the currently circulating variant, but the WT monovalent was not. To account for this, during the Omicron wave we increased the Omicron monovalent efficacy by 10 percentage points (the change in protection for all groups when moving from the Omicron wave to Omicron*) and bivalent protection by 5 percentage points (half of what was applied for Omicron monovalent, because it contains both WT and Omicron) (Figure S6).

### Relative benefit

Equation 1 quantifies the difference in booster benefit among different formulations, which we call the relative benefit.

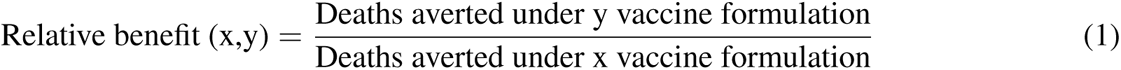

There are three key cases for the relative benefit:

(i) If (and only if) [Booster y deaths averted] *<* [Monovalent (WT) deaths averted], then the relative benefit of booster y is less than 1.0.
(ii) If (and only if) [Booster y deaths averted] *>* [Monovalent (WT) deaths averted] and ([Booster y deaths averted]*-* [Monovalent (WT) deaths averted]) *<* [Monovalent (WT) deaths averted], then the relative benefit is between 1.0 and 2.0.
(iii) If (and only if) [Booster y deaths averted] *>* [Monovalent (WT) deaths averted] and ([Booster y deaths averted]*-* [Monovalent (WT) deaths averted]) *>* [Monovalent (WT) deaths averted], then the relative benefit is greater than 2.0.

In words, if the relative benefit is less than 1.0, then there is a net loss in the deaths averted when moving from WT monovalent to Omicron monovalent or bivalent. If the new formulation averts more deaths than monovalent (WT), but the gain moving from no-booster to a monovalent (WT) booster is larger than the gain from monovalent (WT) to a new formulation, then the relative benefit is between 1.0 *-* 2.0. Finally, if the new formulation both averts more deaths than monovalent (WT) and the gain moving from no-booster to a monovalent (WT) booster is smaller than the gain from monovalent (WT) to a new formulation, then the relative benefit is greater than 2.0.

### Hybrid-immunity model (HIM)

#### Model structure

We also implemented a compartmental transmission model in R which extended a published COVID-19 transmission model [41] to account for vaccine-specific (none, vaccinated, vaccinated + boosted), natural (0, 1+ infection), and hybrid immunity (combination). Our SEIR-like model includes six infection compartments: Susceptible *S*, Exposed *E*, Asymptomatic *A*, Symptomatic *I*, Recovered *R*, and Deceased *D* (Figure S5). The model was also stratified by age (*<* 20, 20 *-* 64, and ≥ 65 years), socioeconomic status (high vs. low), prior natural infection (no natural infection or one or more prior infection), and vaccine status (unvaccinated, primary series, and boosted). Susceptible individuals become exposed through contact with an infectious person (asymptomatic or asymptomatic), after which they enter a latent period (in the exposed *E* class). After leaving the latent class, individuals develop either symptomatic or asymptomatic infection. All asymptomatic individuals recover. Some symptomatic individuals recover, but some fraction dies. Those in the low socioeconomic group have higher mortality rates [4], higher transmission rates because of differences in pandemic mobility [4], and slower rates of vaccine rollout [5] than their higher income counterparts. General parameters are shown in Table S3.

#### Natural infection

After recovery, individuals infected for the first time have sterilizing immunity for 300 days (10 months), after which point they enter the susceptible class for individuals with prior infection (*S*_2_). Our main simulations assume that 50% of those who have been infected previously have waned as of October 2022, consistent with most natural immunity having been acquired during the omicron wave, but we consider an alternative where 75% have waned as a sensitivity analysis. Individuals who have been previously infected have a lower rate of infection (1 *-* 0.8*cp*) and mortality (1 *-cp*), which is influenced by the level of cross protection (*cp*) between the prior infecting strains (based on the population-averaged immune history) and the currently circulating strain. Upon re-infection, previously infected individuals can follow the same steps as for the compartments in the base model. We only model two levels of infection, assuming that infectivity and severity are similar for secondary and higher infections. The level of pre-existing immunity and cross protection at the start of the Omicron* wave was estimated for each country based on the population immune history profiles from the history-specific model (Table S9).

#### Vaccination

*Primary series vaccination*. Vaccination was implemented by adding additional compartments for vaccinated/vaccinated and boosted individuals, which mirror the compartments in the base model: *S_V_*, *E_V_*, *A_V_*, *I_V_*, *R_V_*, and *D_V_* for vaccination. We assumed that everyone who was planning to receive their primary vaccine series had already done so as of October 2022, so only booster vaccines were included in our main model simulations, matching the HSM. Baseline vaccine prevalence in October 2022 was set in the same was as the HSM, following published data from [5].

Vaccine efficacy for the primary series was assumed to reduce the risk of mortality by 70%, consistent with data on Astra-Zeneca protection after 6 months [42–45]. While primary series vaccination for Astra-Zeneca can reduce risk of infection by about 50%, these benefits are only present shortly after vaccination [42–45]. Given that the populations included in our model completed their vaccine campaigns earlier and current rollout rates are low, we do not model any protection against infection for the primary vaccination class.

*Booster vaccination*. To capture the potential for booster vaccination, we added additional compartments (*S_B_*, *E_B_*, *A_B_*, *I_B_*, *R_B_*, and *D_B_*). For simplicity, we assume that individuals in the *S*, *E*, or *R* classes can be vaccinated but individuals in the other compartments will not be. To reflect the fact that coverage of vaccine boosters was low in LMICs in October 2022, when HIM simulations began, only one boosted class was used and its efficacy was varied to reflect the potential benefit of variant specific boosters. Booster vaccination moves vaccinated individuals to the corresponding booster class, but does not confer specific temporary immunity (unlike the HSM). In other words, *S_V_* individuals move to the *S_B_* class, not the *R_B_* class. Similar to the HSM, we only model adult boosting. We implemented vaccination using dynamic daily rates using the same approach as the history-specific model (Table S8), where daily doses were administered until coverage saturates at its peak level. We considered an optimistic roll out as our default scenario, where the halfway week of the rollout curve was sped up by 10 weeks compared to primary series vaccination, but also considered a conservative rollout as a sensitivity analysis where the speed of boosting matches primary series vaccination (Table S2).

#### Hybrid immunity

We also explicitly accounted for joint natural immunity and vaccine-derived immunity–individuals who have both exposures are tracked in a vaccinated and infected classes (*S_V_* _2_, *E_V_* _2_, *A_V_* _2_, *I_V_* _2_, *R_V_* _2_, and *D_V_* _2_) or vaccinated and boosted classes (*S_B_*_2_, *E_B_*_2_, *A_B_*_2_, *I_B_*_2_, *R_B_*_2_, and *D_B_*_2_) and their protection from both exposures is modeled as multiplicative. For example, bivalent vaccination reduces risk of infection by 74%. In Malaysia, cross protection from prior infection was estimated at 72.8%. Thus, an individual with both exposures has an infection risk of (1 *-* 0.74) *⇥* (1 *-* 0.73) ⇥ β or a combined reduction in infection risk of 93%. We do not differentiate between the order of vaccination and natural infection in the hybrid immunity model.

Booster vaccination can enhance protection against both infection (*V E_i_*) and severe disease (*V E_h_*). We use scenarios consistent with the individual-based model bivalent booster efficacy (*V E_i_*= 0.74, *V E_h_*= 0.925) and monovalent booster efficacy (*V E_i_* = 0.657, *V E_h_* = 0.5256) in the main analysis. While this yields a lower efficacy against severe disease specifically for the monovalent booster (*V E_h_*) compared with primary series vaccination, the total protection against severe disease is higher (84%) due to the inclusion of protection against infection.

## Data Availability

All data produced in the present study will be provided at a public repository upon publication in a journal.

## Acknowledgements

This work was funded by the World Health Organization. The authors would like to thank the Biocluster at the Carl R. Woese Institute for Genomic Biology, University of Illinois Urbana-Champaign, for providing access to the computing resources.

## Data and materials availability

All codes and data necessary to evaluate the conclusions of the paper and reproduce the figures in the main text will be made available upon publication.

