## Supplementary Materials for "Immune history influences SARS-CoV-2 booster impacts: the role of efficacy and redundancy"

### S1 Supplementary text

#### Population structure

In the HSM, we divided the population of each country into age groups of 0-20, 21-65, and 66+ relative to their true country-specific distributions [33]. These population distributions were scaled to have a total size of approximately 100,000 individuals (Table S4). For the HIM, actual population sizes were used in the simulations with age groups 0-19, 20-64, and 65+. For both models, 50% of each population was assigned to be high and low socioeconomic status (SES) respectively.

#### History specific model

##### Variant-wave transitions

Beginning with Wild Type (WT) SARS-CoV-2 at day 0 of the pandemic, we then simulated the Delta wave (beginning on day 420), Omicron wave (day 630) and a new hypothetical variant Omicron* (day 900) which lasts until the end of the simulations on day 1095 (year 3). Due to the computational intensity of a four-variant, history-specific model as well as limitations in data availability for multi-variant histories, we made a simplifying assumption that each variant wave is discrete (no co-circulation of variants). At the end of a previous wave, when a new wave begins any remaining infected individuals are converted to the new variant. In addition, at the beginning of each wave 25 exposed individuals per SES class are seeded into the model to ensure that immediate die-out does not occur.

##### Force of infection

Each of the first three waves has a unique baseline probability of infection given contact, denoted by $\beta$. This value is also scaled by the agent's prior immune history at 80% of the protection for severe disease that are shown in Figure 1A. To capture relative differences in contact across countries, we started with pre-pandemic estimates of contact for children, adults, and elderly [35] and took weighted averages to get total contact across countries. Because we modeled historical waves, we required wave-specific estimates of the probability of infection given contact, and these were taken from household secondary attack rates [7]. Consequently, to account that our probabilities of infection were taken from close contact, we scaled down total contacts by 75% to get baseline contact values for the model. Because mobility tends to occur more within SES-groups rather than outside of it [36, 37], we assumed 60% of contacts are with individuals from the same SES group and 40% are from the opposite group.

Next, we applied pandemic-related reductions in contact. Social distancing is known to be unequal across SES-groups [4], and we accounted for this relative disparity by giving low SES a 30% social distancing reduction and high SES a 60% reduction within each country (Table S5). Finally, each country has a varied history of NPIs including masking or stay-at-home orders. To capture the relative sizes of each wave, reflected in historical confirmed cases for each country (Figure S1), we parameterized a stringency index from 0-1 that applies to both socioeconomic groups within a country, that reduces the effective contact for each wave (Table S1). During the transition from Omicron to Omicron*, stringency is held constant, but we consider the probability of becoming infected to be 10% or 30% higher, representing an increase in infectiousness of the pathogen.

#### Death or recovery

The probability of death following infection, denoted by $\alpha$, is stratified by age and socioeconomic status using previous estimates by our group [4], and scaled by immune history (Figure 1A). If an individual recovers instead, the variant they were infected with is appended to their immune history. We assume that peak immunity, whether natural or vaccine-induced, lasts 10 months.

#### Waned immunity

Ten months after an immune event, immunity wanes to a lower level and agents move to the susceptible class. The degree to which their protection from infection and severe disease is dampened depends on how many immune events $n$ they have previously had, with more prior events $n$ reducing the amount of waning – if $n = 1$, protection wanes to 40% of initial peak protection, if $n = 2$ then 70%, and if $n \geq 3$ then 85%.

#### Vaccines and boosting

Using country- and SES-specific vaccination rates over time previously published by our group [5], we incorporated an initial vaccine intervention during the WT, Delta, and Omicron waves, considering the shape of initial rollout ($k$), eventual maximum covered ($V_m$) and week at which half of the population is vaccinated ($W_h$) (Table S2). The vaccine intervention starts at day 320, representing that vaccination started several months before Delta became the prevailing strain, and continues until the end of the Omicron wave. Anyone who is not currently infectious and has not previously received a vaccine is eligible to be vaccinated, and vaccination moves the agent to the recovered class, with vaccination appended to their immune history. At each time step, the model computes the proportion of the total population that should be vaccinated that day, and attempts to draw that number of individuals from the vaccine-eligible pool. If the eligible pool is smaller than the prescribed number to be vaccinated, the model vaccinates all individuals in the eligible pool.

Starting on day 840 (two months before Omicron*) and continuing until the end of the simulations, we implemented a booster intervention. Maximum potential booster uptake ($B_m$)for each country was based on uptake for the previous campaign of monovalent boosters [39], with the assumption that only vaccinated adults are eligible. To get these coverages, we first calculated the proportion of the total vaccinated individuals that belong to each SES class, based on the maximum vaccinated values for each country (Table S2). We then calculated the number of these doses which went to adults, using the population structure of each country (Table S4). Assuming that boosters would continue to have the same distribution across low and high SES as for the primary series vaccines, we calculated the maximum number of booster doses given to each SES class, then divided these values over the number of vaccinated adults to parameterize the peak coverage among vaccinated adults (Table S8). We used the same shape ($kB$) as the primary series vaccination. We considered two speeds, where timing ($WB_h$) was the same (supplementary analysis) or 10 weeks faster (main text) than primary series vaccination (Figure S3).

#### Booster protection scenarios

The precise responses to booster vaccines stratified by strain-specific immune history, are presently unknown. For this reason, we considered several hypotheses of booster efficacy during Omicron*, using proportional changes deduced from the known immune response to monovalent primary series vaccines during the WT wave. We considered bivalent and WT monovalent boosters, as well as a hypothetical Omicron monovalent booster. Previous work suggests that monovalent boosters may have 40% of the efficacy of bivalent boosters against severe infection [40]. Informed by this estimate, we assumed that a bivalent formulation would have the same numeric change in protection that was originally conferred by primary series vaccination during the WT wave, while monovalent boosters confer 40% of this original numeric change.

##### Scenario 1: History Dependent

For WT monovalent boosters, each individual's numeric change in protection is 40% of their numeric change in protection during primary series vaccination against WT. We also constructed hypothetical Omicron monovalent boosters, where the response of 'WT + vaccine' and 'Omicron(*) + vaccine' are flipped. In the bivalent case, WT and Omicron(*) histories receive a response equal to the average of the WT and Omicron(*) responses during the WT wave, because the booster contains both WT and Omicron antigens. There is no data on primary series vaccination for individuals with an Omicron* history since this is a hypothetical scenario, and thus their responses cannot be individually calculated. Consequently, for the purposes of vaccination and boosting, we assume that individuals who have been exposed to the new Omicron* variant respond the same to each booster as those with Omicron histories.

##### Scenario 2: Same Efficacy

In this scenario, individuals receive a numeric change in protection equal (bivalent) or 40% less (monovalent) to the average observed change across 'WT + vaccine', 'Delta + vaccine', 'Omicron + vaccine', and 'Vaccine' histories during primary series vaccination against WT.

##### Scenario 3: Same Endpoint

In this scenario, we move all individuals to the same end protection parameter, equal to the maximum protection observed in the same efficacy scenario.

##### Scenario 4: No boosting

Finally, we considered a scenario in which no booster is implemented.

##### Calculating events

We used a modified tau-leap algorithm to determine which events (infection, recovery, etc.) would occur and when [34]. Vaccination and boosting proceeds separately from the tau-leap, and is performed at time intervals of 1 day.

#### Hybrid immunity model

##### Model structure

The flow of individuals through the model was as follows. Initially, many individuals are susceptible to infection (S). Upon exposure, they enter a latent period (E), during which they cannot transmit. They can then develop asymptomatic infection (entering the A class) or symptomatic infection (entering the I class). We assume that all asymptomatic individuals will recover (R). Those with symptomatic infections can either recover (entering the R class) or die (entering the deceased class D), and the rest will recover (entering the R class).

#### Immunity

For country-specific benchmarking for the HIM, we calculated the weighted average of cross protection among existing prior infections prior to boosting (Table S9). This was calculated by combining the wave specific cross protection for each natural infection history against Omicron* and multiplying by their relative abundance among the previously infected population.

#### Force of infection

As a starting point, we used contact matrices estimated for each country pre-pandemic, stratified by age [35]. The probability of infection given contact among all contacts was calibrated to match the force of infection in the HSM (Table S3). We also tested two scenarios where infectiousness is not reduced by pre-existing immunity: one where the probability of infection is unchanged, and another where the probabilities of infection were recalibrated at 4.79% for low SES and 1.82% for high SES to still match the infectiousness of Omicron [23]. While the HIM included assortatitivity in contacts by age, it did not include assortativity by SES, though overall contact was lower for high SES matching the HSM. We also used results from the HSM calibration to parameterize the baseline stringency for each site so that our force of infection values were relatively comparable.

#### Booster coverage, efficacy, and rollout rates

Since the HIM starts at the start of the Omicron* wave and the main simulations assumed that boosting started 60 days before Omicron*, we calculated the starting booster coverages using the type-III functional response functions with the expected rollout curve from days 0 to 60 and using day 60 as initial coverage on day 1 of the HIM simulations. Rollout rates were parameterized to match the HSM, including the peak booster coverages adjusted to exclude children and unvaccinated from boosting (Table S8). Because immune history was not explicitly modeled and was captured using baseline immunity and cross protection due to natural infection, the HIM roughly corresponds to the same endpoint or same efficacy scenarios from the individual based model (Figures S11, S10.

#### Initial conditions

The initial age distribution of infections and deaths were estimated using seroprevalence data and incidence data from Our World In Data (OWID) [46]. To initialize the model, we calculated a reporting rate by comparing seroprevalence data reported from the individual countries with reported cases from Our World In Data as of 12 days before the estimated seroprevalence was conducted to allow time for seroconversion [47–49]. The estimated reporting rates for both symptomatic and asymptomatic infections were 0.178 for Malaysia, 0.053 for India, and 0.189 for Ecuador. We assumed that only symptomatic infections were reported. Reported cases as of October 12, 2022 were multiplied by 1/reporting rate to get the total number of active symptomatic infections on that date. We assumed that only 40% of infections were symptomatic, so asymptomatic infections were assumed to be 1.5 times the total active symptomatic conditions. Initial latent infections were set equal to I(0)+E(0).

For our main simulations, we set baseline immunity for the country-specific simulations to its estimated value from the HSM. Then, to explore how future dynamics for any country depend on prior infection, we varied the level of baseline immunity from 0-100%. For each level of immunity, we assumed that the prevalence of immunity was the same by vaccine strata and 50% of previously infected cases were assumed to have waned immunity into the higher S class. For example, for a scenario with 50% baseline immunity, 25% of the unvaccinated population was put into the $R$ class and 25% of this class was put into $S_2$. No individuals were assumed to be in the $R_2$ classes at the start of the simulation, equivalent to assuming that no individuals had experienced more than 1 prior infection. We also relaxed this assumption in sensitivity analyses, and the results were similar.

### S2 Supplementary Figures

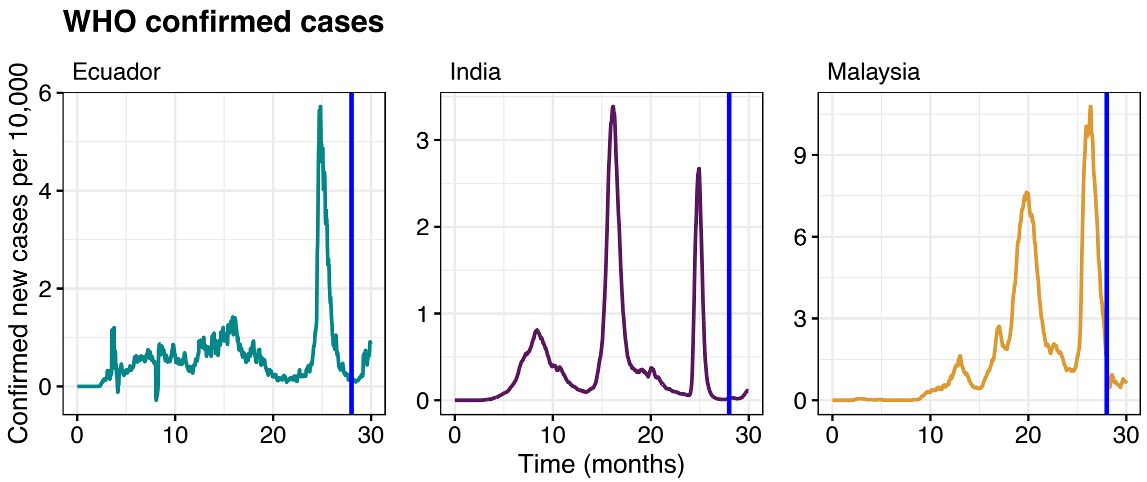


Figure S1: Moving 7-day average of confirmed cases per 10,000 population prior to the Omicron* wave (data from WHO [21]). Blue lines denote the start of booster vaccination in the HSM.

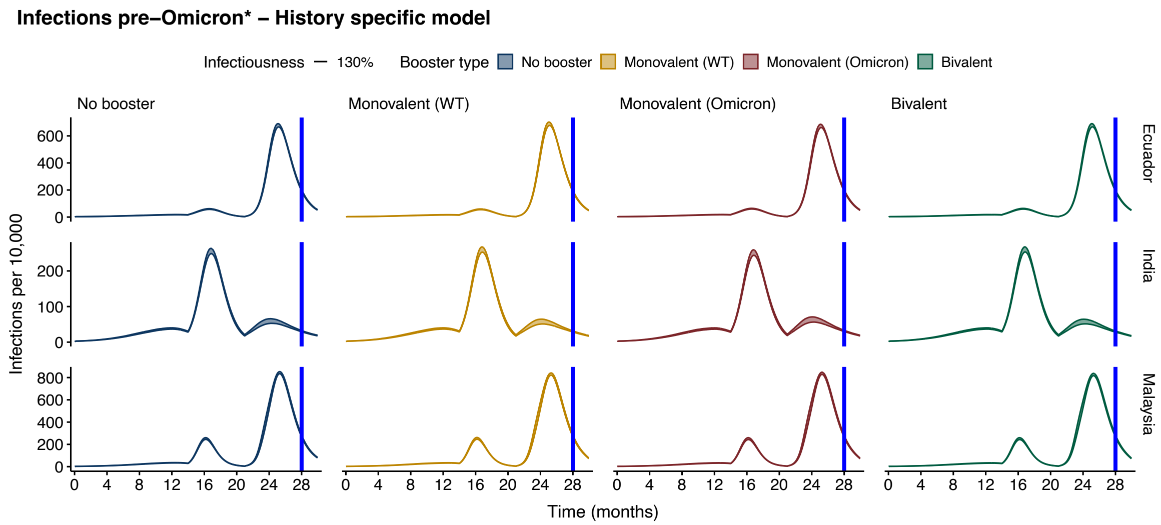


Figure S2: HSM-simulated infections before Omicron*, with 500 replicates for each scenario. 95% confidence intervals from the t-distribution are shown (ribbons). Blue lines denote the start of booster vaccination in the HSM.

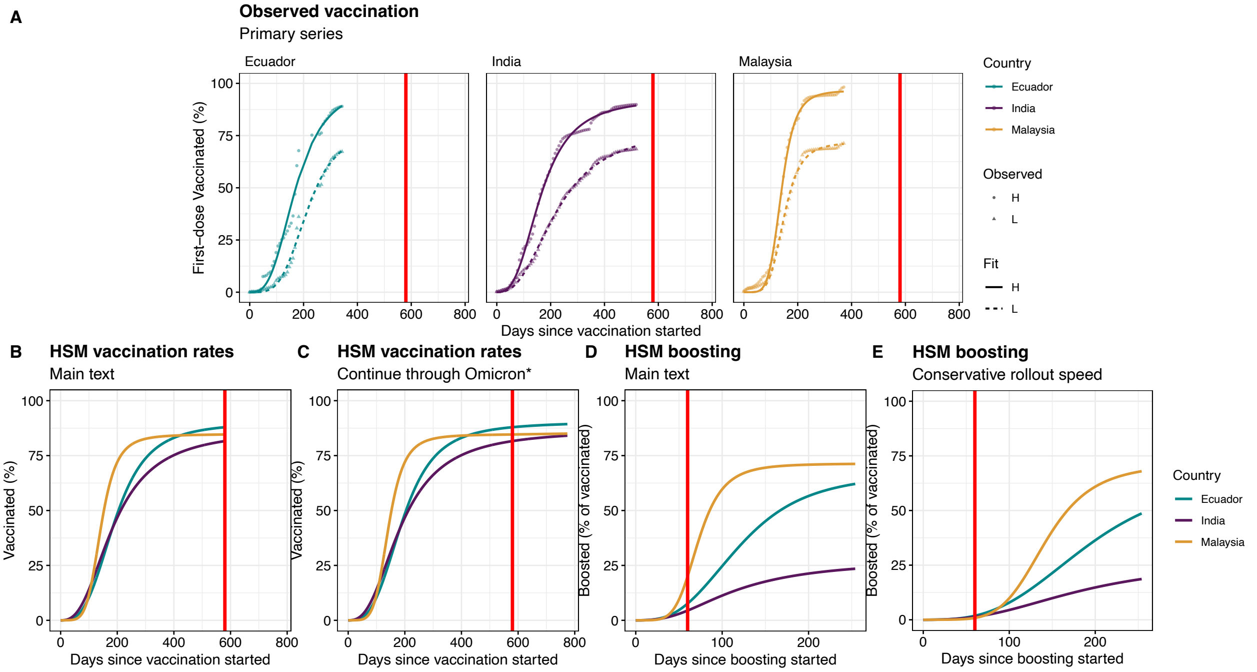


Figure S3: HSM vaccine and booster trends. (A) Observed and fit data of primary series vaccination for Ecuador, India, and Malaysia [5]. (B) Vaccination rates in the HSM, if vaccination stops at the end of the Omicron wave. (C) Vaccination rates in the HSM, if vaccination continues through Omicron*. (D) Booster rates in the HSM, if booster timing is 10 weeks faster than primary series vaccination for all groups. (E) Booster rates in the HSM, if booster timing is the same as primary series vaccination. Red lines denote the start of Omicron*.

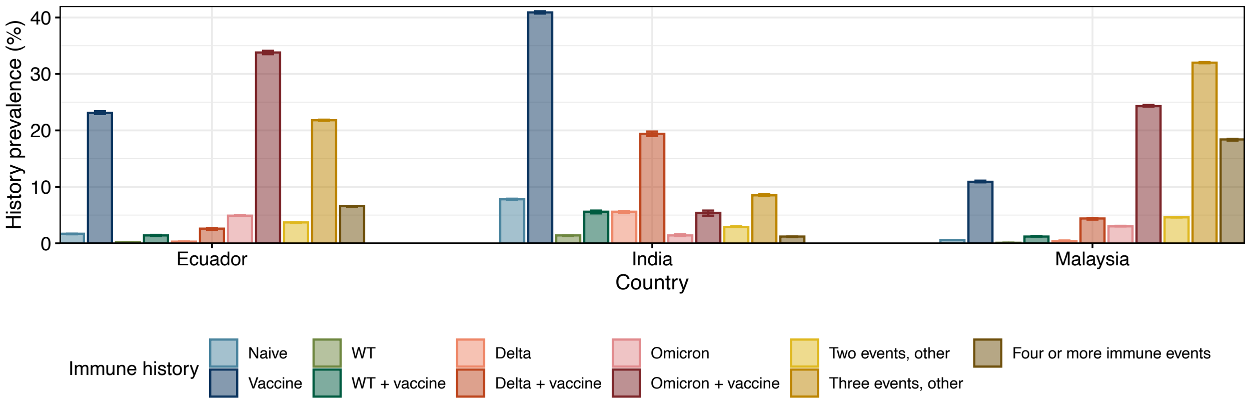


Figure S4: Distribution of immune histories by country in the HSM on day 839 with 500 replicates, as seen in Figure 1A, prior to the beginning of booster vaccination. 95% confidence intervals from t-distribution are shown (whiskers).

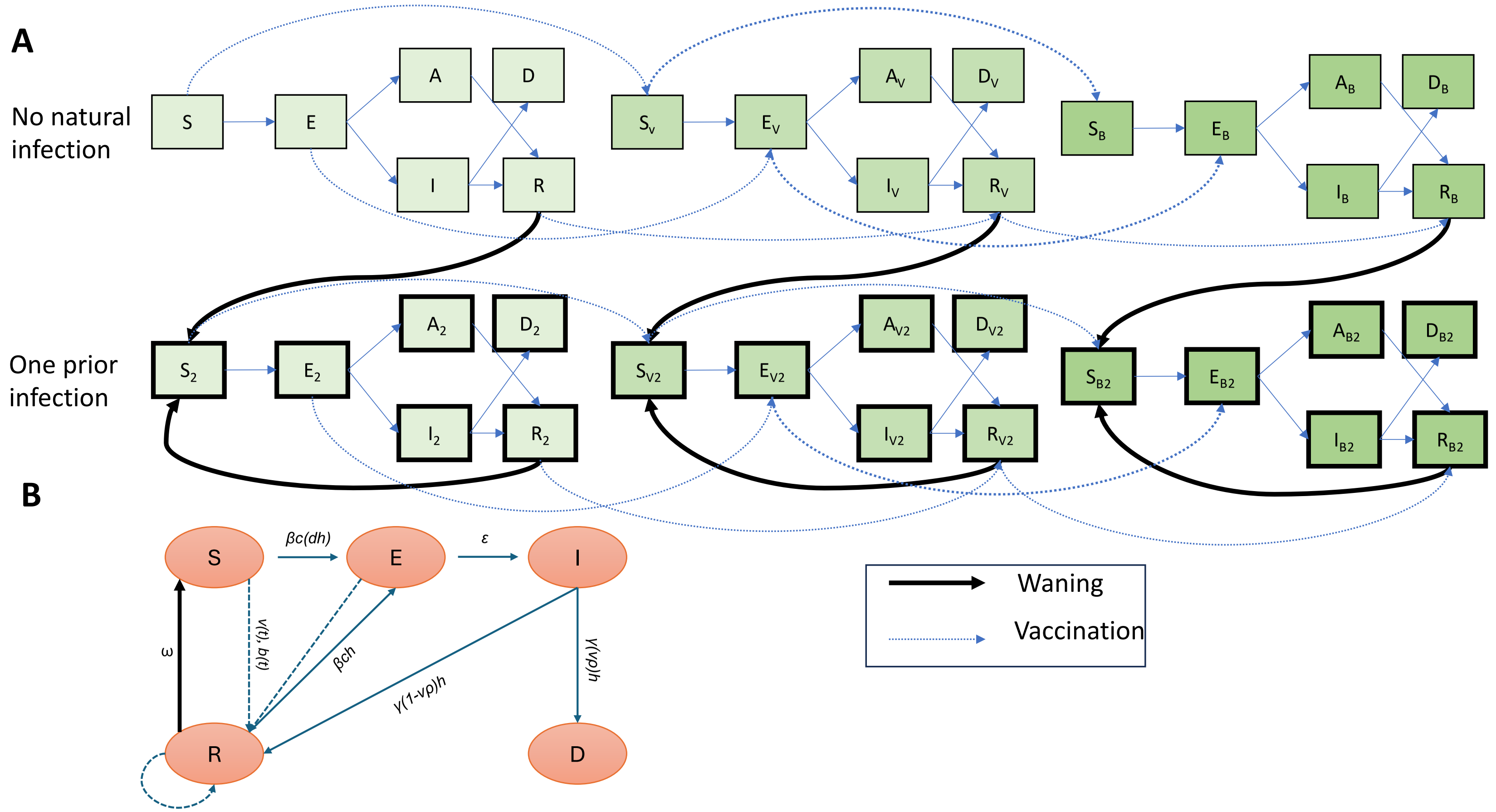


Figure S5: Compartmental model diagram for the hybrid immunity model (A) and history specific model (B). In panel A, box colors indicate vaccine status (lightest green: unvaccinated, medium green: primary series vaccination, dark green: boosted). For both models, the dark black line indicates waning immunity and the dotted line shows vaccination events. In panel (B), parameter $h$ represents the immune history scaling parameter for infections (when multiplied by $\beta$) or deaths (when multiplied by $\rho$).

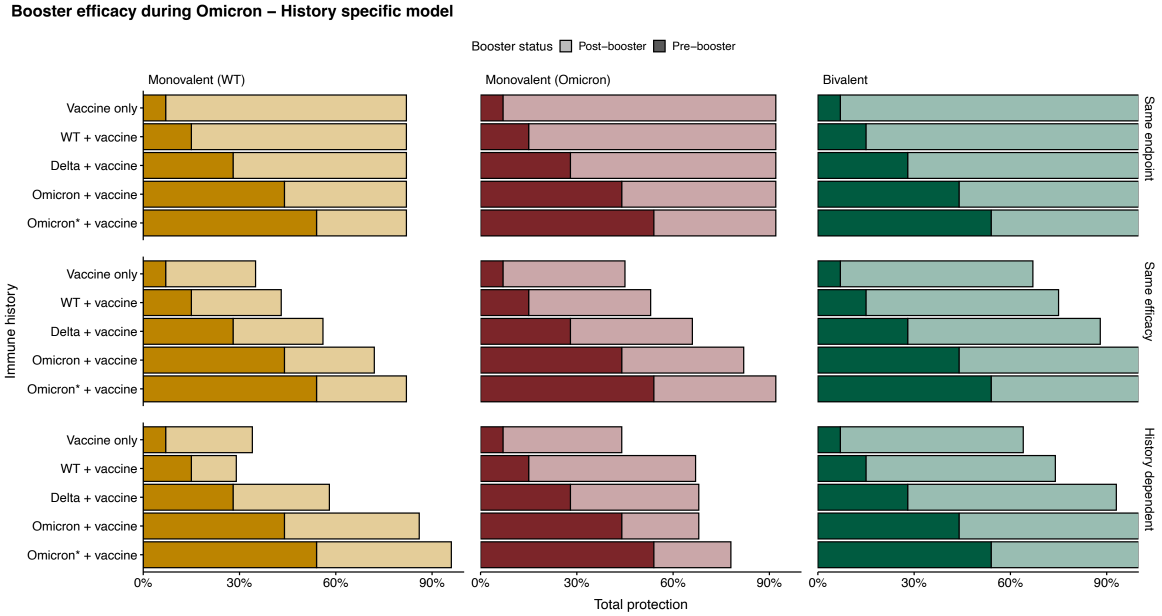


Figure S6: Booster protection against severe disease during the Omicron wave by immune history, under a bivalent, WT monovalent, or hypothetical Omicron monovalent formulation. These values are used for 60 days, from the start of boosting at day 840 until the start of the Omicron* wave on day 900.

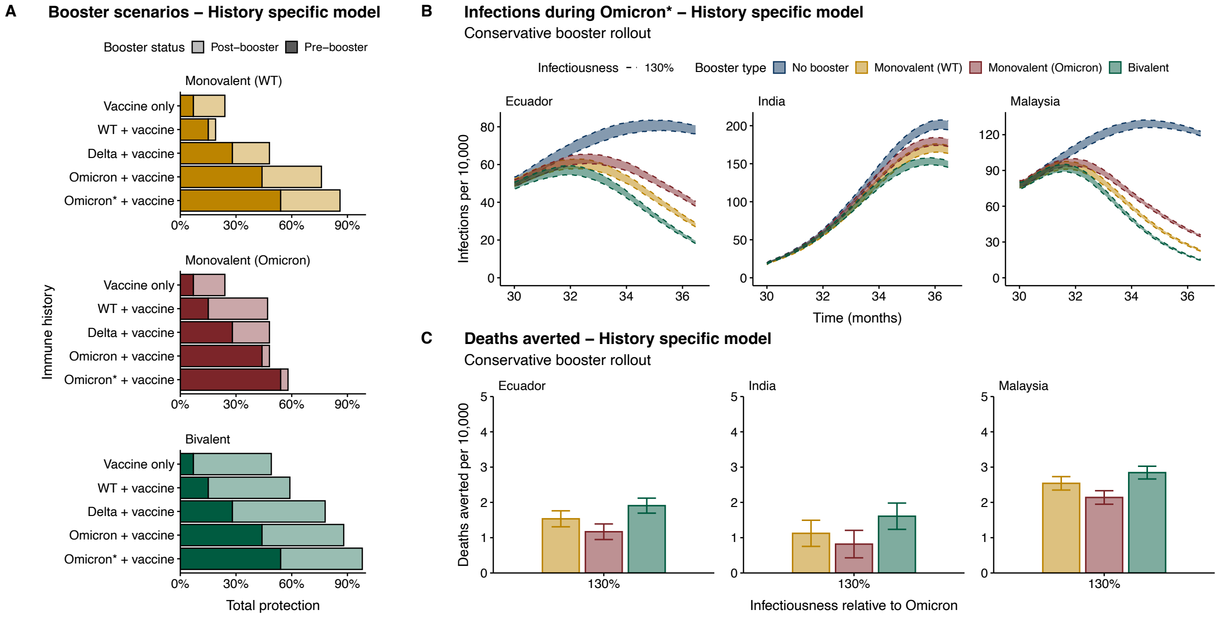


Figure S7: HSM booster impacts under the assumption that the timing of booster vaccination is the same as primary series rollout. (A) Booster protection against severe disease during the Omicron* wave, by immune history, under a bivalent, WT monovalent, or hypothetical Omicron monovalent formulation. (B) Infection trends with 500 replicates under three boosters or a no-boosting scenario for Ecuador, India, and Malaysia if Omicron* is 10% or 30% more infectious than Omicron. 95% confidence intervals from the t-distribution are shown (ribbons). (E) Deaths averted by boosting since the start of Omicron* (30 months) through the end of simulations (36.5 months), with 500 replicates under each booster. 95% confidence intervals from the t-distribution are shown with whiskers. The HSM model structure is shown in Figure S5. General simulation parameters are shown in Table S3, with country-specific population structure in Table S4, contact rates by country and SES in Table S5, wave- and country-specific stringency in Table S1, and vaccination parameters by country and SES in Table S2. The maximum boosted and shape of boosting parameters are shown in Table S8, and the timing of boosting matches primary series vaccination (Table S2).

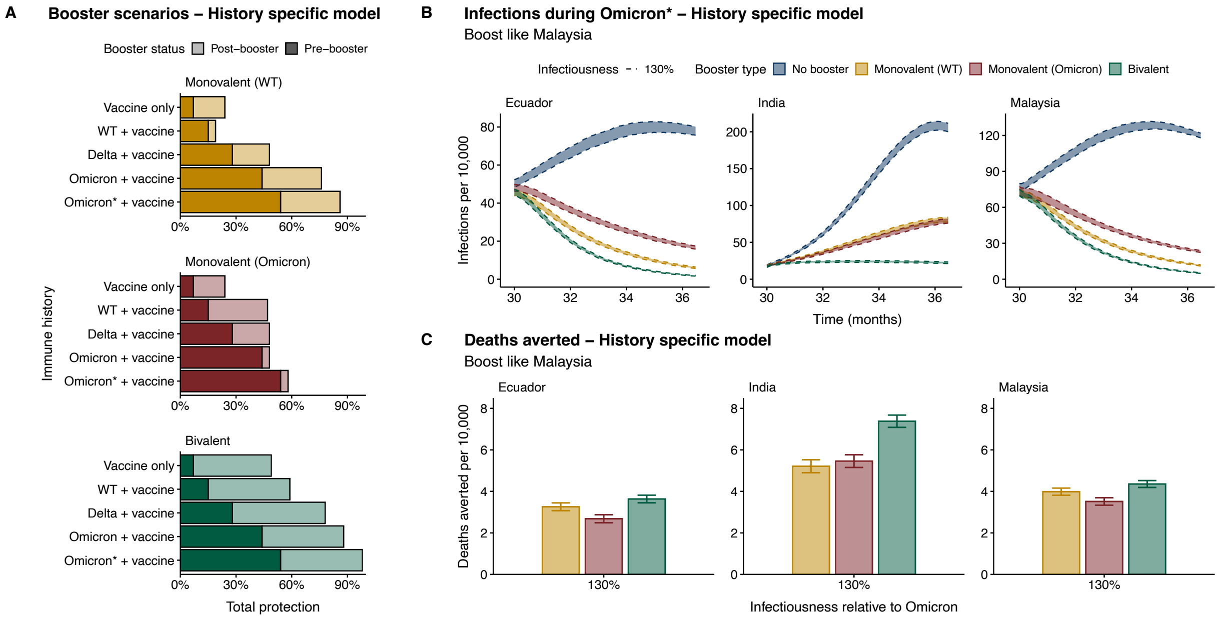


Figure S8: HSM booster impacts under the assumption that all countries boost with the same booster trend as Malaysia. (A) Booster protection against severe disease during the Omicron* wave, by immune history, under a bivalent, WT monovalent, or hypothetical Omicron monovalent formulation. (B) Infection trends with 500 replicates under three boosters or a no-boosting scenario for Ecuador, India, and Malaysia if Omicron* is 10% or 30% more infectious than Omicron. 95% confidence intervals from the t-distribution are shown (ribbons). (E) Deaths averted by boosting since the start of Omicron* (30 months) through the end of simulations (36.5 months), with 500 replicates under each booster. 95% confidence intervals from the t-distribution are shown with whiskers. The HSM model structure is shown in Figure S5. General simulation parameters are shown in Table S3, with country-specific population structure in Table S4, contact rates by country and SES in Table S5, wave- and country-specific stringency in Table S1, and vaccination parameters by country and SES in Table S2. Boosting parameters for Malaysia are shown in Table S8.

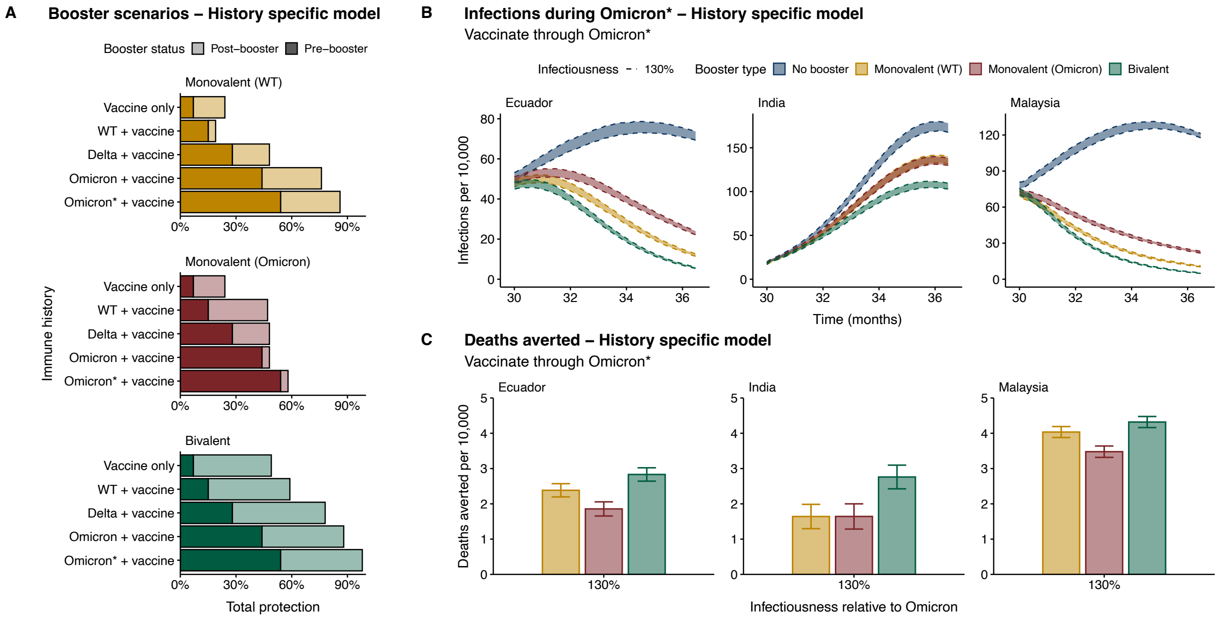


Figure S9: HSM booster impacts under the assumption that vaccination continues through Omicron*. (A) Booster protection against severe disease during the Omicron* wave, by immune history, under a bivalent, WT monovalent, or hypothetical Omicron monovalent formulation. (B) Infection trends with 500 replicates under three boosters or a no-boosting scenario for Ecuador, India, and Malaysia if Omicron* is 10% or 30% more infectious than Omicron. 95% confidence intervals from the t-distribution are shown (ribbons). (E) Deaths averted by boosting since the start of Omicron* (30 months) through the end of simulations (36.5 months), with 500 replicates under each booster. 95% confidence intervals from the t-distribution are shown with whiskers. The HSM model structure is shown in Figure S5. General simulation parameters are shown in Table S3, with country-specific population structure in Table S4, contact rates by country and SES in Table S5, wave- and country-specific stringency in Table S1, and vaccination parameters by country and SES in Table S2. Boosting parameters are shown in Table S8.

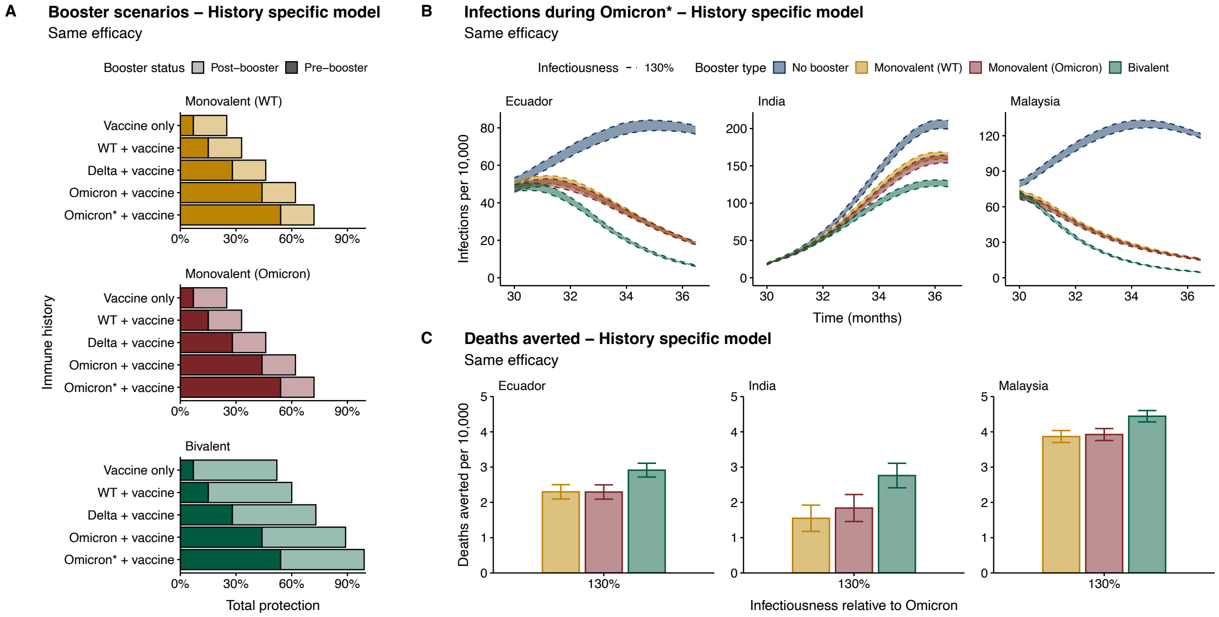


Figure S10: HSM booster impacts under the assumption that boosting results in the same numeric change in protection for all individuals ("Same efficacy"). (A) Booster protection against severe disease during the Omicron* wave, by immune history, under a bivalent, WT monovalent, or hypothetical Omicron monovalent formulation. (B) Infection trends with 500 replicates under three boosters or a no-boosting scenario for Ecuador, India, and Malaysia if Omicron* is 10% or 30% more infectious than Omicron. 95% confidence intervals from the t-distribution are shown (ribbons). (E) Deaths averted by boosting since the start of Omicron* (30 months) through the end of simulations (36.5 months), with 500 replicates under each booster. 95% confidence intervals from the t-distribution are shown with whiskers. The HSM model structure is shown in Figure S5. General simulation parameters are shown in Table S3, with country-specific population structure in Table S4, contact rates by country and SES in Table S5, wave- and country-specific stringency in Table S1, and vaccination parameters by country and SES in Table S2. Boosting parameters, which are assumed to be 10 weeks faster than for primary series vaccination, are shown in Table S8.

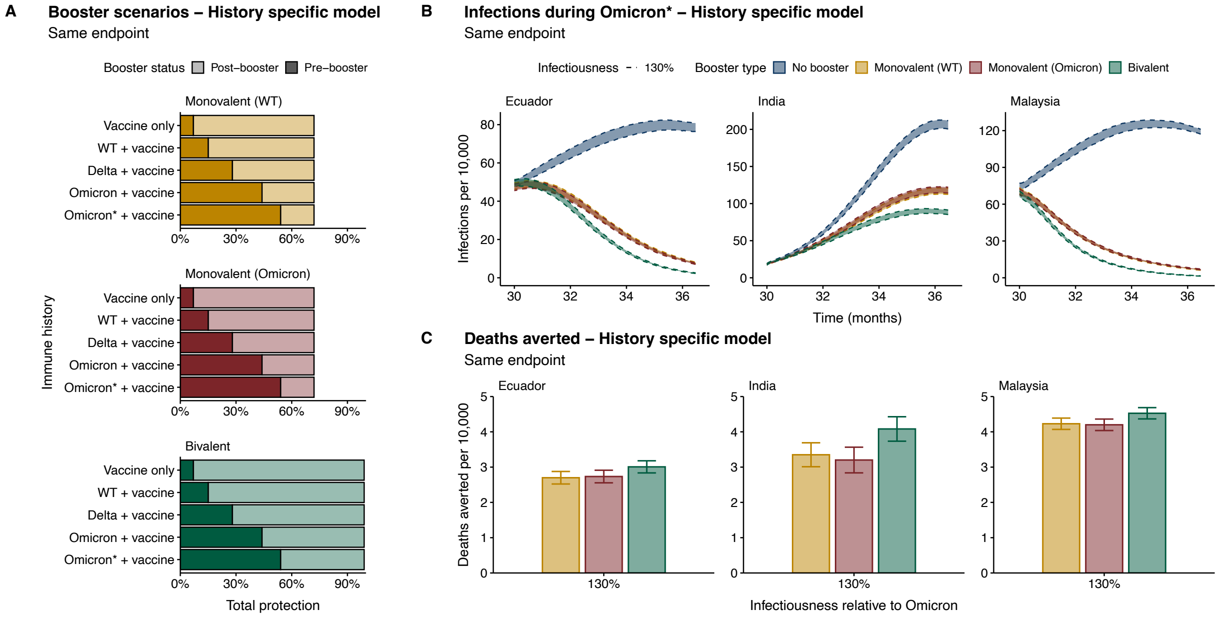


Figure S11: HSM booster impacts under the assumption that boosting results in the same end protection for all individuals (”Same endpoint”). (A) Booster protection against severe disease during the Omicron* wave, by immune history, under a bivalent, WT monovalent, or hypothetical Omicron monovalent formulation. (B) Infection trends with 500 replicates under three boosters or a no-boosting scenario for Ecuador, India, and Malaysia if Omicron* is 10% or 30% more infectious than Omicron. 95% confidence intervals from the t-distribution are shown (ribbons). (E) Deaths averted by boosting since the start of Omicron* (30 months) through the end of simulations (36.5 months), with 500 replicates under each booster. 95% confidence intervals from the t-distribution are shown with whiskers. The HSM model structure is shown in Figure S5. General simulation parameters are shown in Table S3, with country-specific population structure in Table S4, contact rates by country and SES in Table S5, wave- and country-specific stringency in Table S1, and vaccination parameters by country and SES in Table S2. Boosting parameters, which are assumed to be 10 weeks faster than for primary series vaccination, are shown in Table S8.

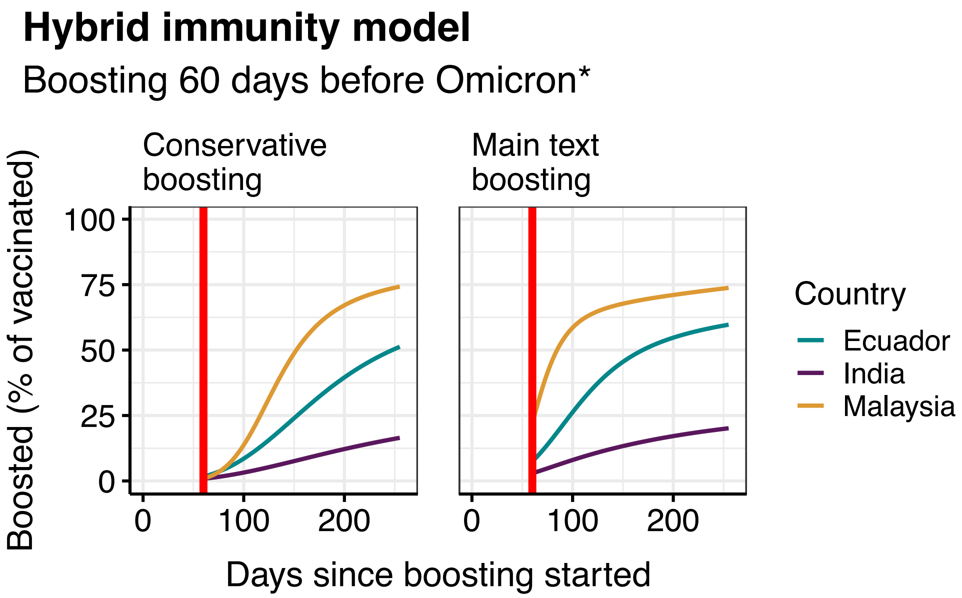


Figure S12: HIM booster curves. Percentage of vaccinated adults boosted over time, under conservative or main-text booster rollout, if boosting starts 60 days prior to Omicron*. Red lines denote the start of Omicron*.

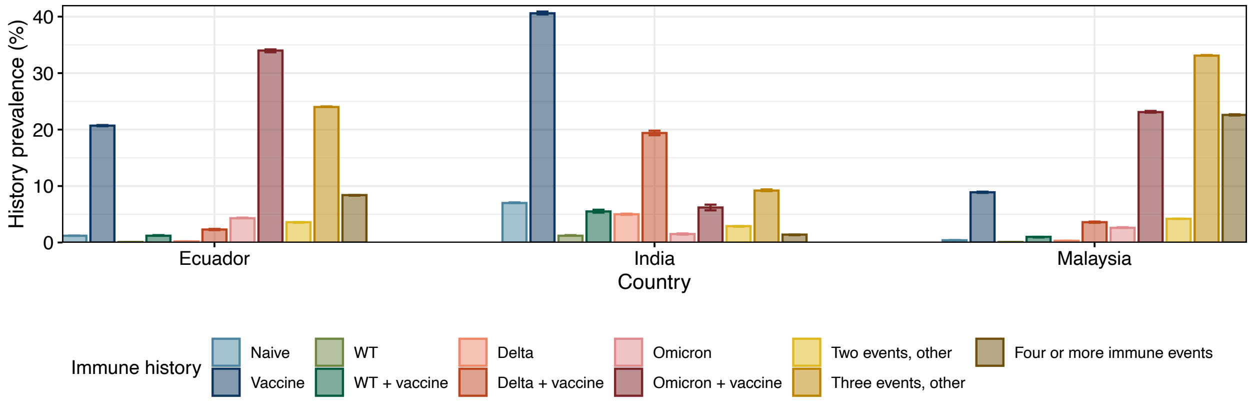


Figure S13: Distribution of immune histories by country in the HSM on day 899, under a no booster scenario with 500 replicates - used to estimate cross protection and number of prior infections in the HIM (Table S9). 95% confidence intervals from t-distribution are shown (whiskers).

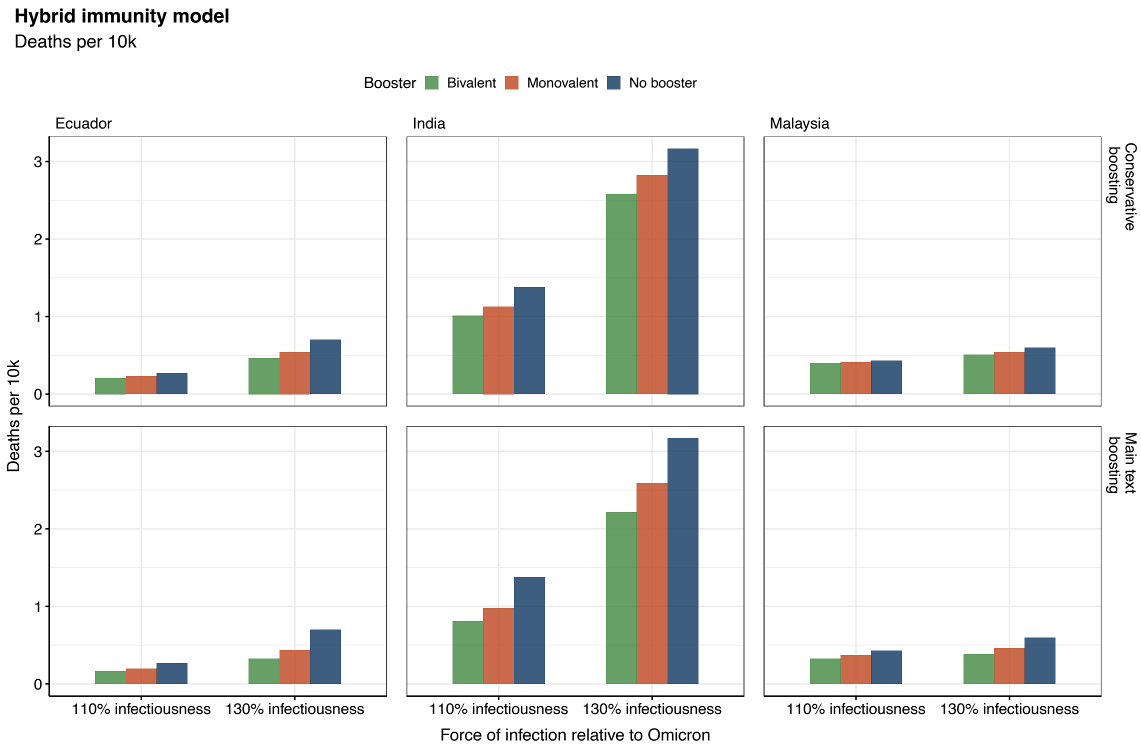


Figure S14: Total deaths per 10,000 in the HIM, during Omicron*, if booster rollout is conservative or follows main-text speed. The HIM model structure is shown in Figure S5. General parameters are shown in Table S3, stringency in Table S1, initial conditions in Table S10, cross-protection in Table S9, and boosting parameters in Table S8. In the conservative booster scenario, booster rollout speed $WB_h$ matches the timing of primary series vaccination (Table S2).

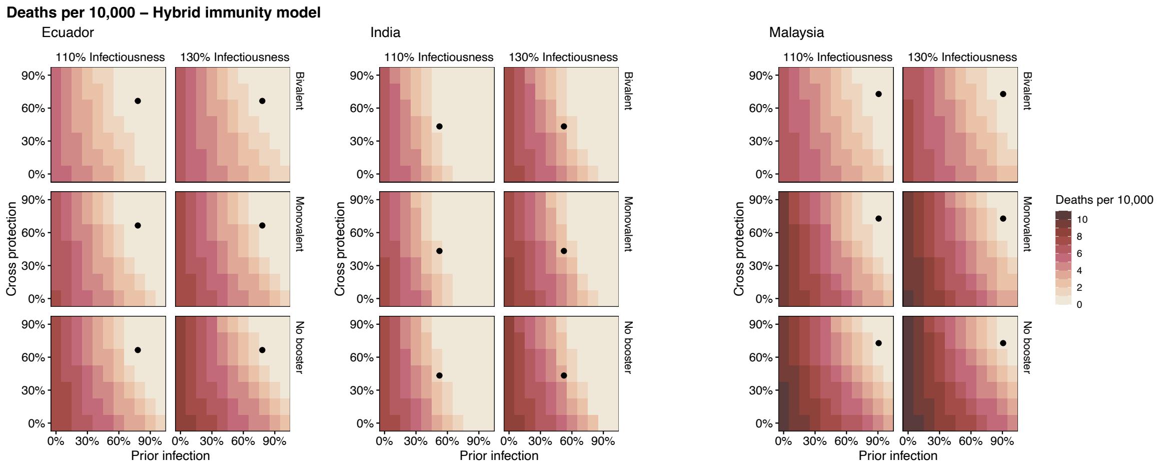


Figure S15: Total deaths per 10,000 in the HIM, during Omicron*, corresponding to main text scenarios. The HIM model structure is shown in Figure S5. General parameters are shown in Table S3, stringency in Table S1, initial conditions in Table S10, cross-protection in Table S9, and boosting parameters in Table S8.

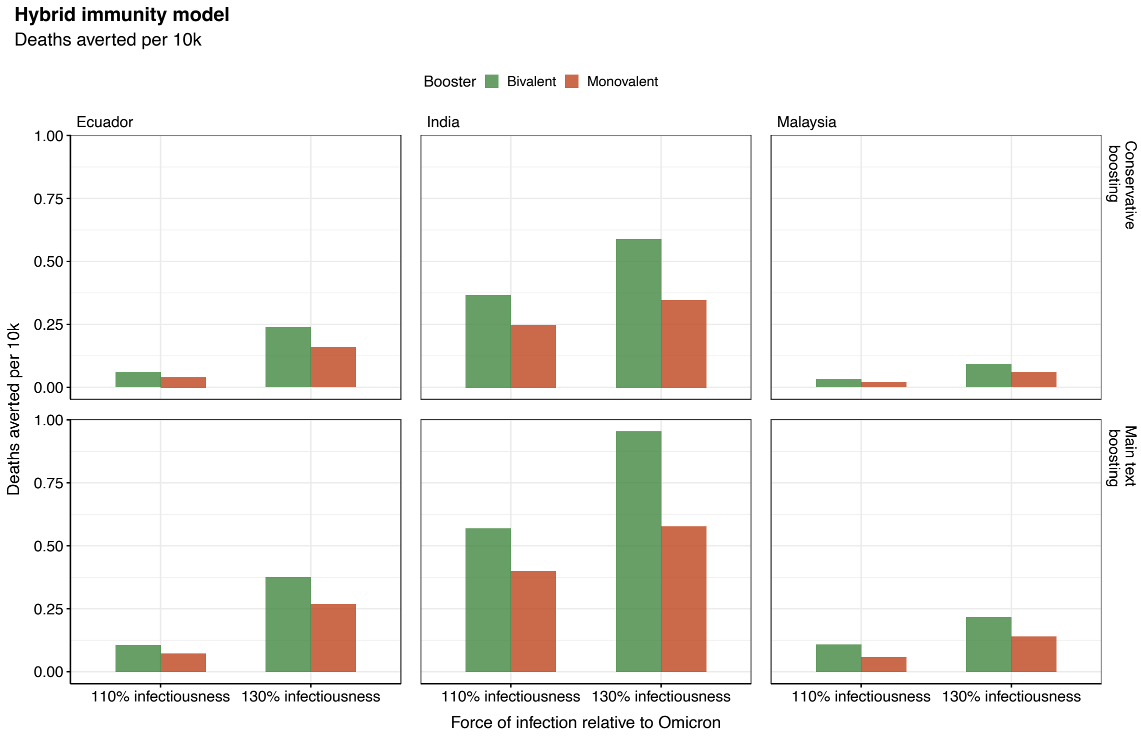


Figure S16: Deaths averted per 10,000 in the HIM during Omicron*, under conservative or main-text booster rollout, if boosting starts 60 days prior to Omicron*. The HIM model structure is shown in Figure S5. General parameters are shown in Table S3, stringency in Table S1, initial conditions in Table S10, cross-protection in Table S9, and boosting parameters in Table S8. In the conservative booster scenario, booster rollout speed $WB_h$ matches the timing of primary series vaccination (Table S2).

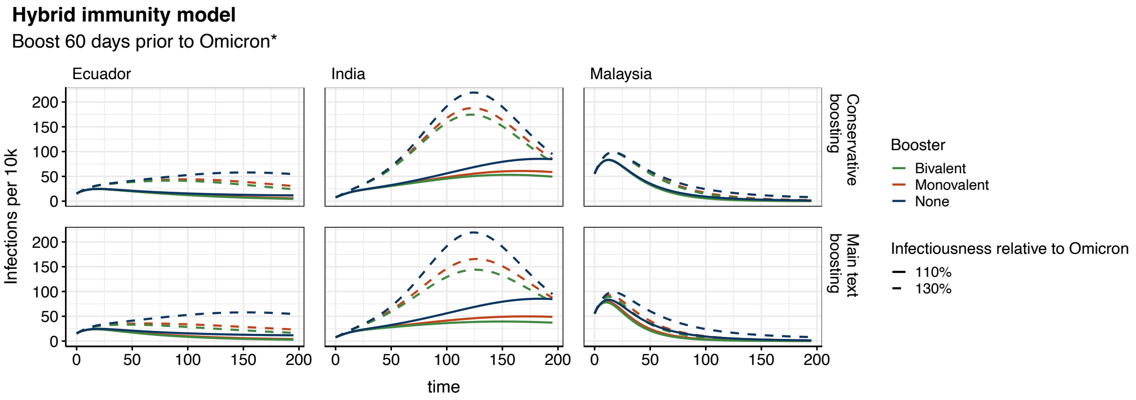


Figure S17: Cases per 10,000 in the HIM during Omicron* under conservative or main-text booster rollout, if boosting starts 60 days prior to Omicron*. The HIM model structure is shown in Figure S5. General parameters are shown in Table S3, stringency in Table S1, initial conditions in Table S10, cross-protection in Table S9, and boosting parameters in Table S8. In the conservative booster scenario, booster rollout speed $WB_h$ matches the timing of primary series vaccination (Table S2).

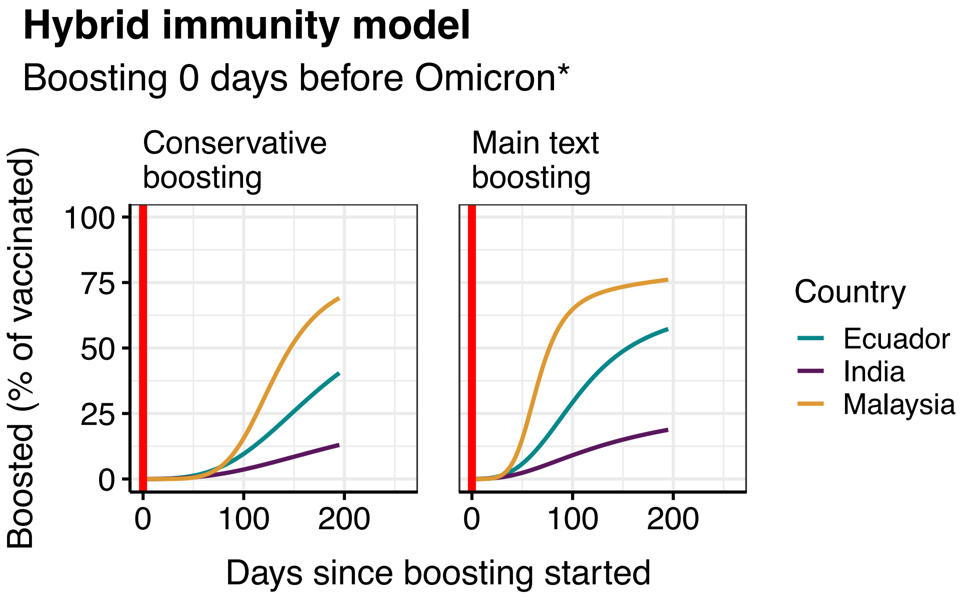


Figure S18: HIM booster curves. Percentage of vaccinated adults boosted over time, under conservative or main-text booster rollout, if boosting starts at the same time as Omicron*. Red lines denote the start of the Omicron* wave.

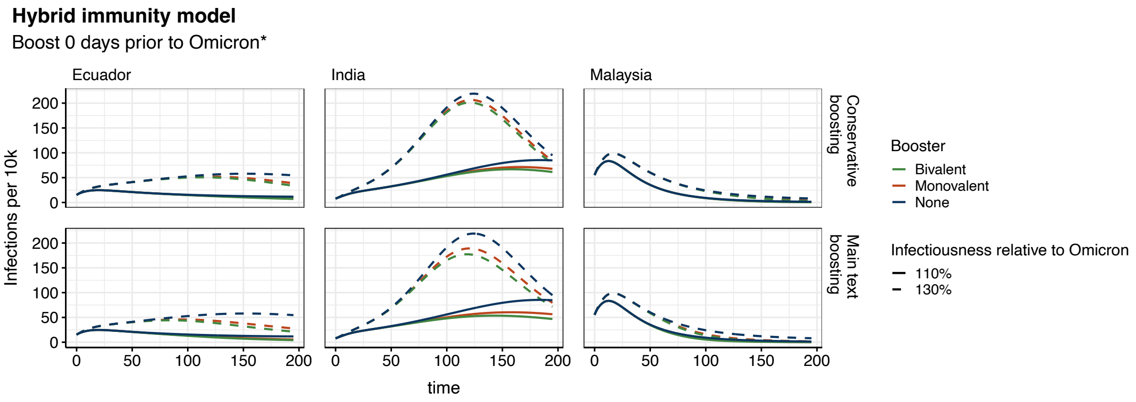


Figure S19: Cases per 10,000 in the HIM during the Omicron* wave, under conservative or main-text booster rollout, if boosting starts at the same time as Omicron*. The HIM model structure is shown in Figure S5. General parameters are shown in Table S3, stringency in Table S1, initial conditions in Table S10, cross-protection in Table S9, and boosting parameters in Table S8. In the conservative booster scenario, booster rollout speed $WB_h$ matches the timing of primary series vaccination (Table S2).

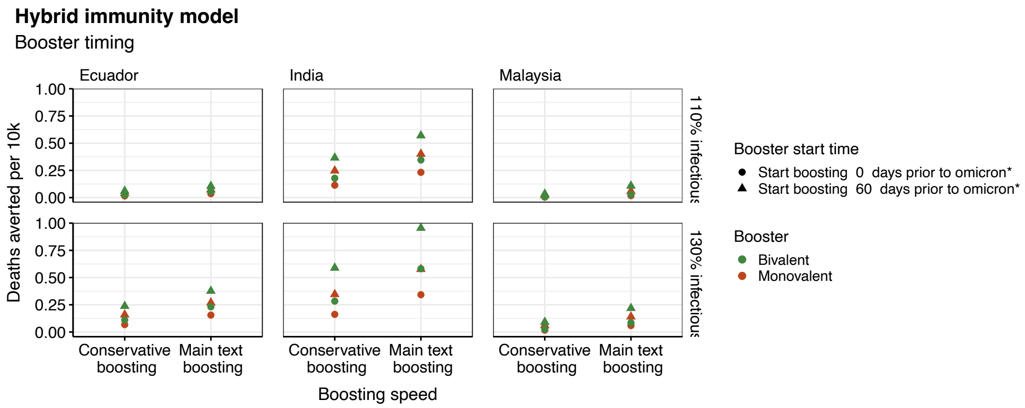


Figure S20: Deaths averted per 10,000 population in the HIM during Omicron*, under conservative or main-text booster rollout. Boosting starts either at the start of Omicron* (circles) or 60 days prior (triangles). The HIM model structure is shown in Figure S5. General parameters are shown in Table S3, stringency in Table S1, initial conditions in Table S10, cross-protection in Table S9, and boosting parameters in Table S8. In the conservative booster scenario, booster rollout speed $WB_h$ matches the timing of primary series vaccination (Table S2).

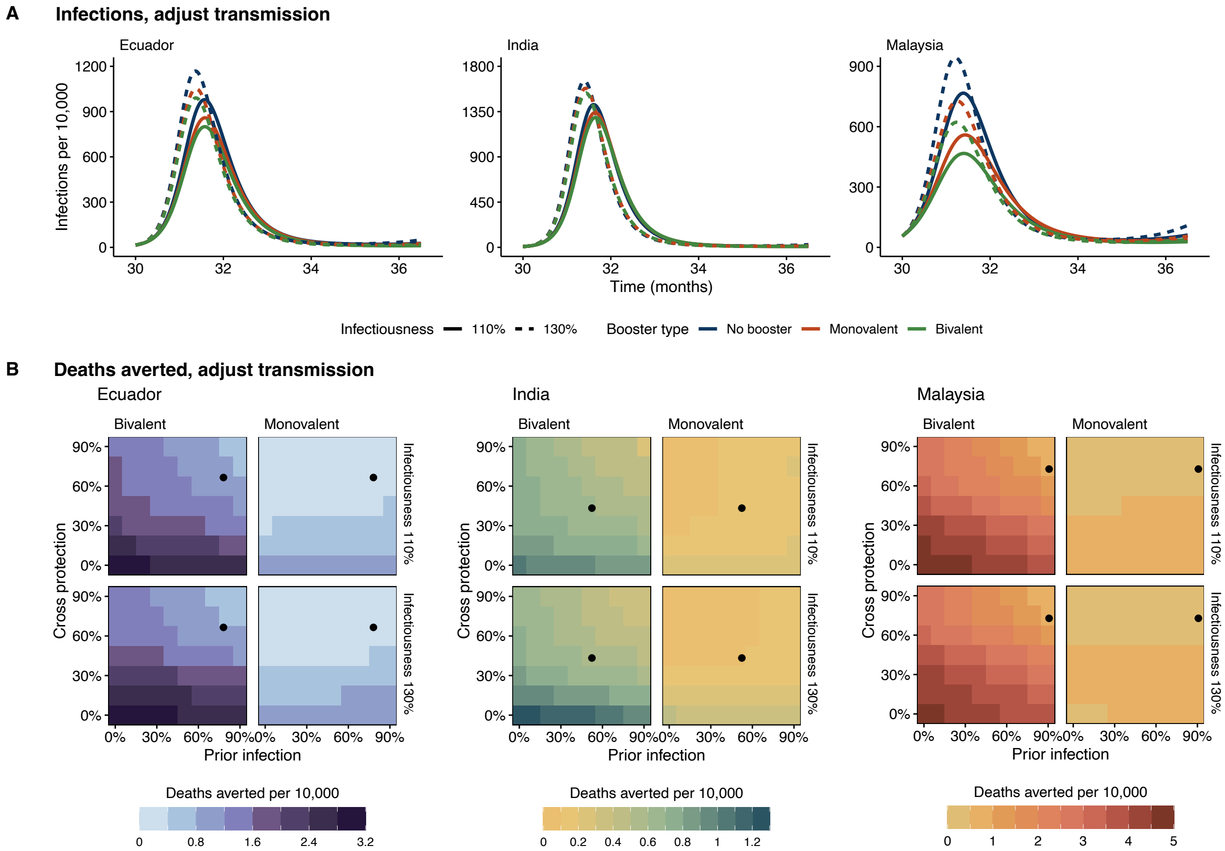


Figure S21: HIM projections, where the assumption that prior immunity reduces transmission, is lifted, but $\beta$ is not recalibrated. (A) Infections per 10,000 during the Omicron* period, where Omicron* is considered to be 10% or 30% more infectious than Omicron. (B) Deaths averted per 10,000 under a bivalent or monovalent booster, compared to a ”no boosting” scenario. Prior immunity represents the percentage of the population that has been previously infected. Cross protection represents the overlap between the population’s immune history and the currently circulating variant. Country-specific immunity levels are included (black dots). The HIM model structure is shown in Figure S5. General parameters are shown in Table S3, stringency in Table S1, initial conditions in Table S10, cross-protection in Table S9, and boosting parameters in Table S8.

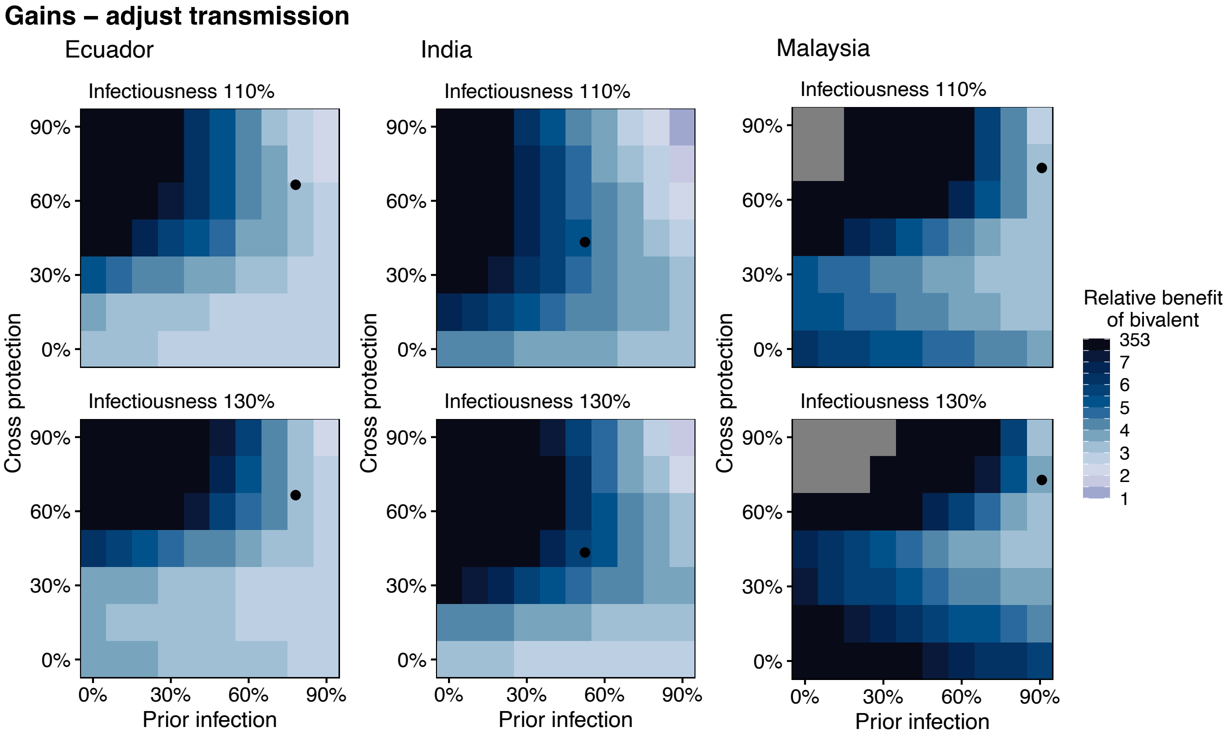


Figure S22: Comparison of bivalent vs. monovalent boosters in the HIM, where the assumption that prior immunity reduces transmission, is lifted, but $\beta$ is not recalibrated. Relative benefit of the bivalent booster is calculated with Equation 1, matching the HSM. Grey values represent areas where zero deaths were averted by a monovalent booster and thus Equation 1 cannot be calculated. Prior immunity represents the percentage of the population that has been previously infected. Cross protection represents the overlap between the population's immune history and the currently circulating variant. Country-specific immunity levels are included (black dots). The HIM model structure is shown in Figure S5. General parameters are shown in Table S3, stringency in Table S1, initial conditions in Table S10, cross-protection in Table S9, and boosting parameters in Table S8.

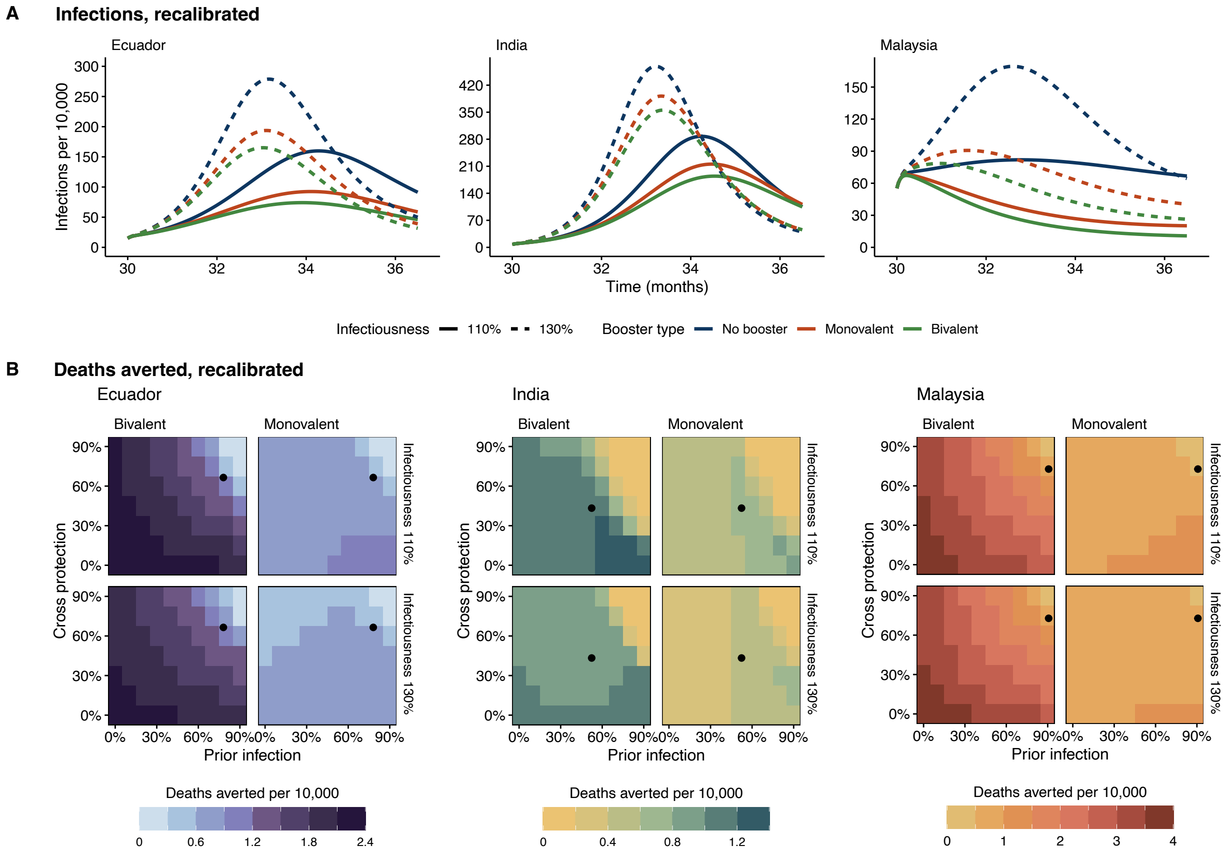


Figure S23: HIM projections, where the assumption that prior immunity reduces transmission, is lifted, and $\beta$ is recalibrated. (A) Infections per 10,000 during the Omicron* period, where Omicron* is considered to be 10% or 30% more infectious than Omicron. (B) Deaths averted per 10,000 under a bivalent or monovalent booster, compared to a ”no boosting” scenario. Prior immunity represents the percentage of the population that has been previously infected. Cross protection represents the overlap between the population's immune history and the currently circulating variant. Country-specific immunity levels are included (black dots). The HIM model structure is shown in Figure S5. General parameters are shown in Table S3, stringency in Table S1, initial conditions in Table S10, cross-protection in Table S9, and boosting parameters in Table S8.

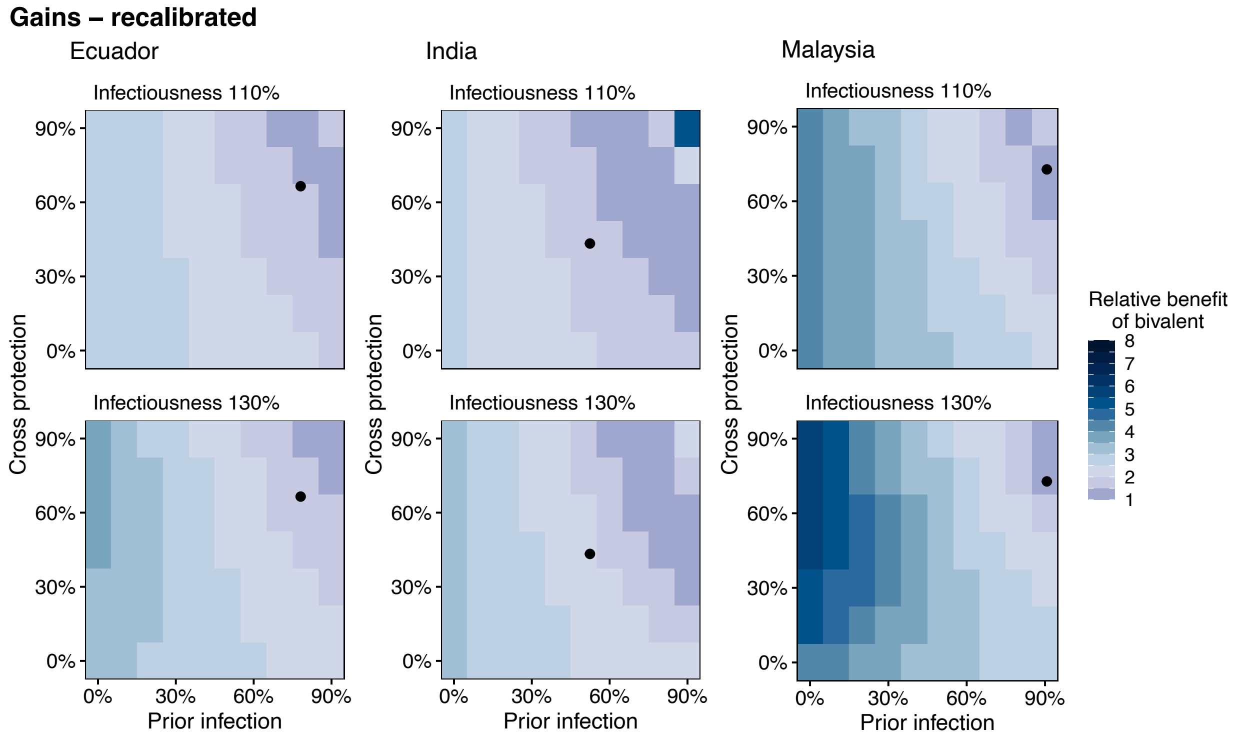


Figure S24: Comparison of bivalent vs. monovalent boosters in the HIM, where the assumption that prior immunity reduces transmission, is lifted, and $\beta$ is recalibrated. Relative benefit of the bivalent booster is calculated with Equation 1, matching the HSM. Prior immunity represents the percentage of the population that has been previously infected. Cross protection represents the overlap between the population's immune history and the currently circulating variant. Country-specific immunity levels are included (black dots). The HIM model structure is shown in Figure S5. General parameters are shown in Table S3, stringency in Table S1, initial conditions in Table S10, cross-protection in Table S9, and boosting parameters in Table S8.

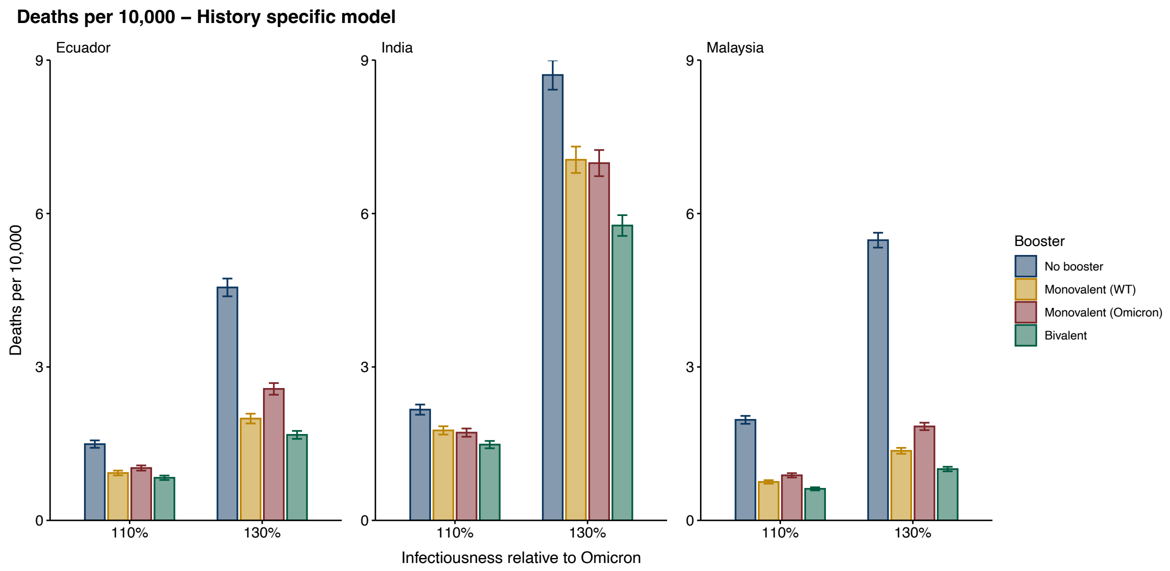


Figure S25: Deaths per 10,000 individuals in the HSM, with 500 replicates per scenario. 95% confidence intervals from t-distribution are shown (whiskers).

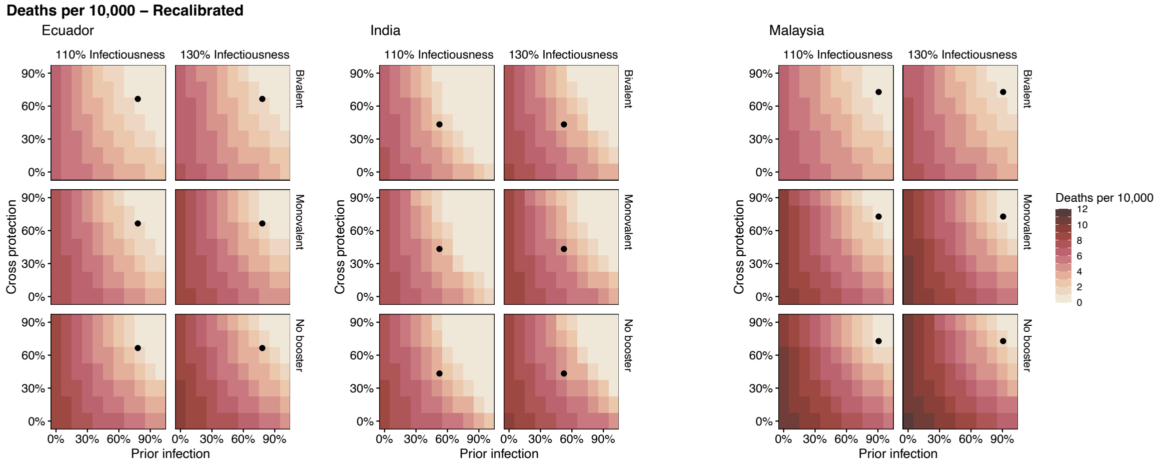


Figure S26: Total deaths per 10,000 in the HIM if the assumption that prior immunity reduces transmission is lifted and $\beta$ is recalibrated. The HIM model structure is shown in Figure S5. General parameters are shown in Table S3, stringency in Table S1, initial conditions in Table S10, cross-protection in Table S9, and boosting parameters in Table S8.

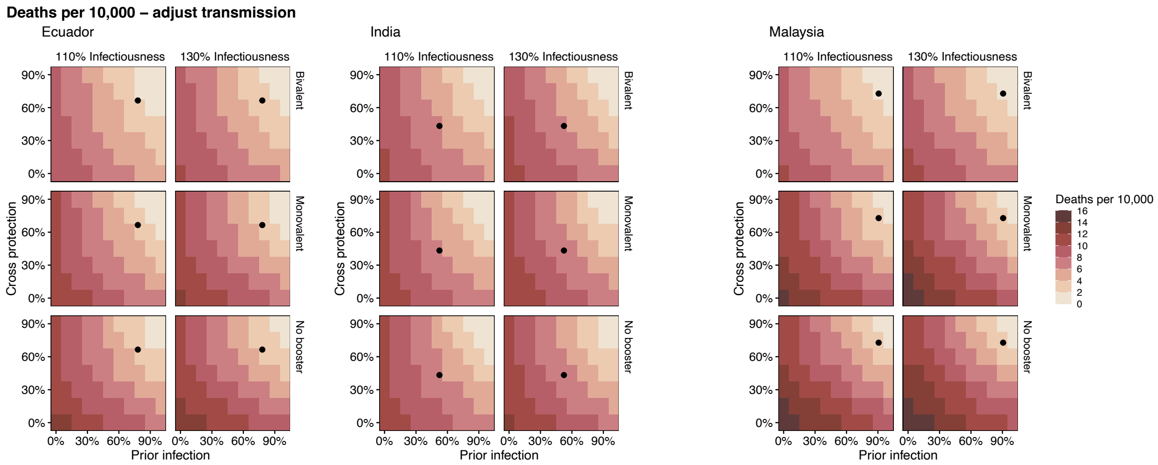


Figure S27: Total deaths per 10,000 in the HIM if the assumption that prior immunity reduces transmission is lifted but $\beta$ is not recalibrated. The HIM model structure is shown in Figure S5. General parameters are shown in Table S3, stringency in Table S1, initial conditions in Table S10, cross-protection in Table S9, and boosting parameters in Table S8.

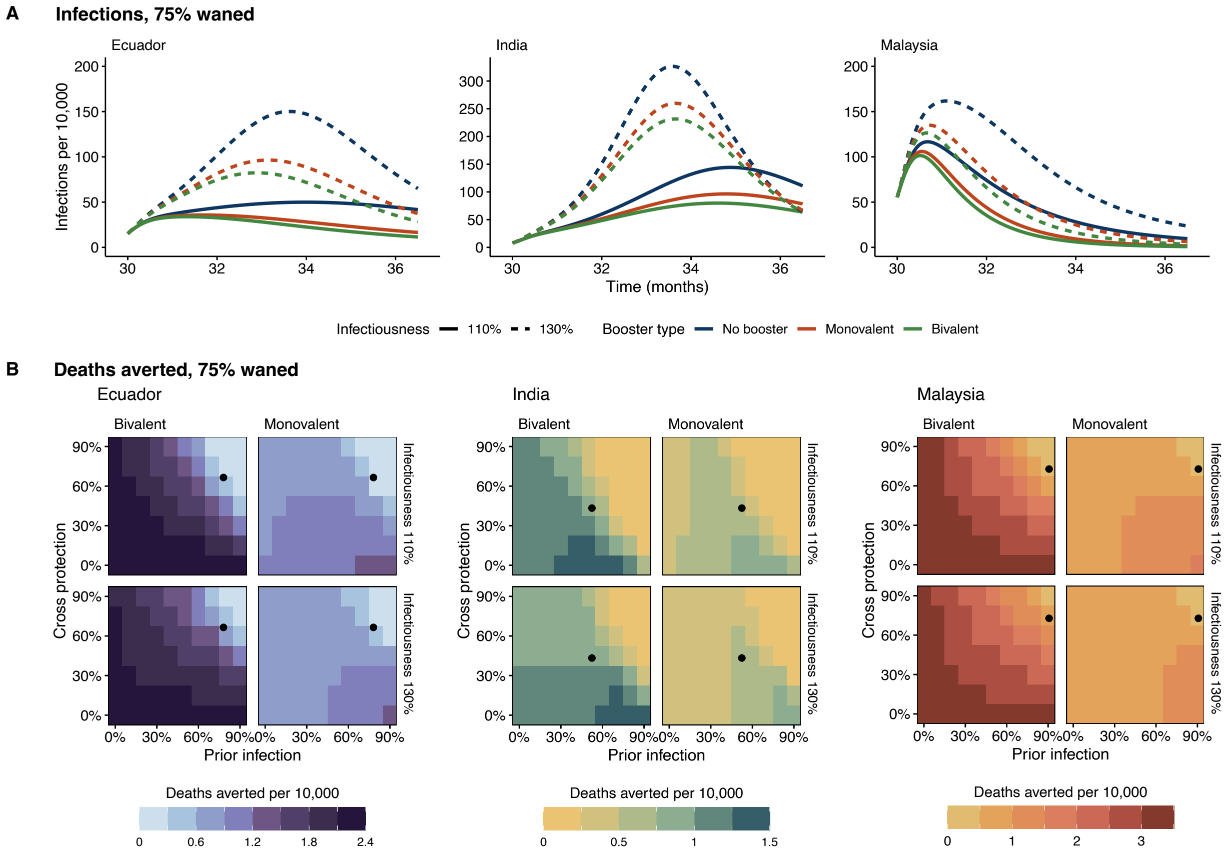


Figure S28: HIM projections, where 75% of previously infected individuals have waned at the start of simulations. (A) Infections per 10,000 during the Omicron* period, where Omicron* is considered to be 10% or 30% more infectious than Omicron. (B) Deaths averted per 10,000 under a bivalent or monovalent booster, compared to a "no boosting" scenario. Prior immunity represents the percentage of the population that has been previously infected. Cross protection represents the overlap between the population's immune history and the currently circulating variant. Country-specific immunity levels are included (black dots). The HIM model structure is shown in Figure S5. General parameters are shown in Table S3, stringency in Table S1, initial conditions in Table S10, cross-protection in Table S9, and boosting parameters in Table S8.

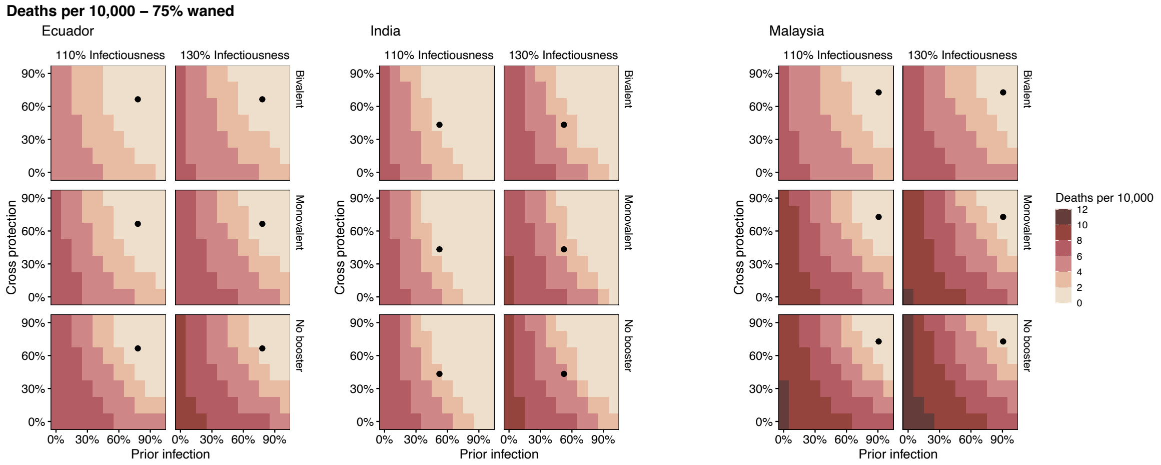


Figure S29: Total deaths per 10,000 in the HIM if 75% of previously infected individuals have waned. The HIM model structure is shown in Figure S5. General parameters are shown in Table S3, stringency in Table S1, initial conditions in Table S10, cross-protection in Table S9, and boosting parameters in Table S8. In the conservative booster scenario, booster rollout speed $WB_h$ matches the timing of primary series vaccination (Table S2).

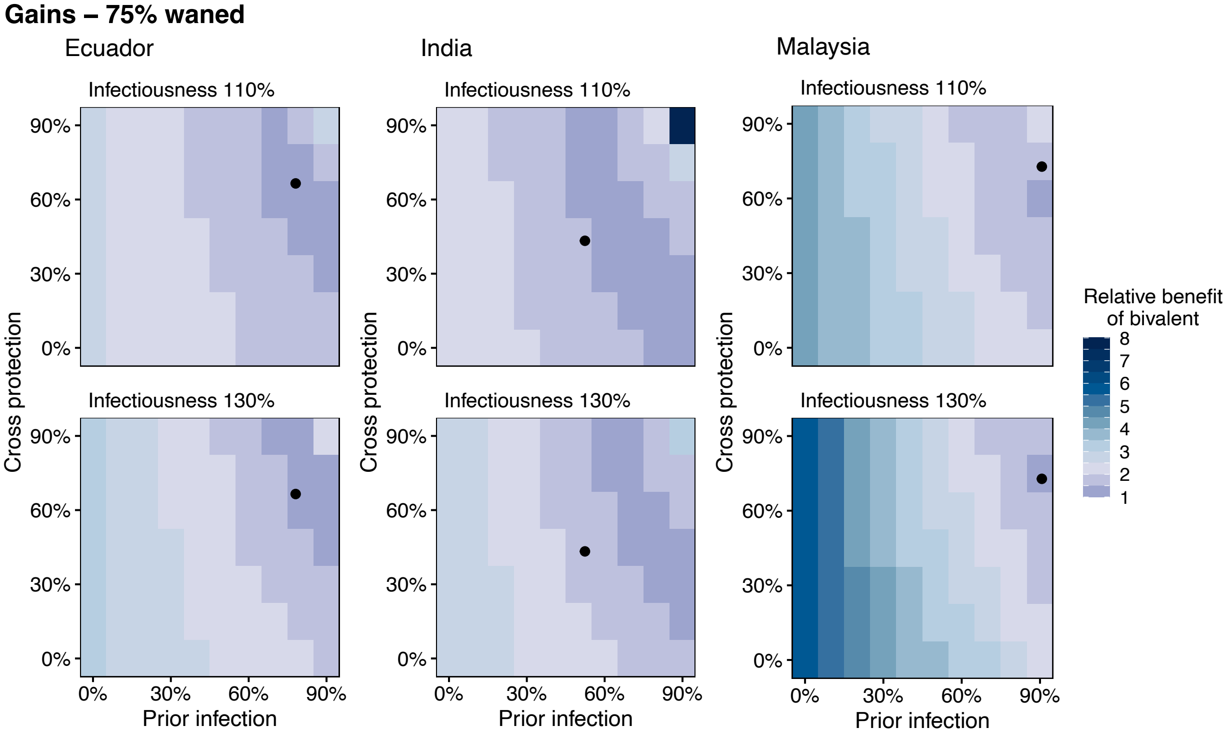


Figure S30: Comparison of bivalent vs. monovalent boosters in the HIM, where 75% of previously infected individuals have waned at the start of simulations. Relative benefit of the bivalent booster is calculated with Equation 1, matching the HSM. Prior immunity represents the percentage of the population that has been previously infected. Cross protection represents the overlap between the population's immune history and the currently circulating variant. Country-specific immunity levels are included (black dots). The HIM model structure is shown in Figure S5. General parameters are shown in Table S3, stringency in Table S1, initial conditions in Table S10, cross-protection in Table S9, and boosting parameters in Table S8.

### S3 Supplementary tables

| Wave | Malaysia | Ecuador | India |
|---|---|---|---|
| WT | 0.42 | 0.40 | 0.22 |
| Delta | 0.51 | 0.35 | 0.24 |
| Omicron | 0.93 | 0.71 | 0.29 |
| Omicron* | 0.93 | 0.71 | 0.29 |

Table S1: Wave-specific stringency across countries.

| Parameter | Meaning | Malaysia | Ecuador | India |
|---|---|---|---|---|
| $k_L$ | Shape of vaccination trends, low SES | 5.0 | 3.9 | 2.5 |
| $k_H$ | Shape of vaccination trends, high SES | 5.5 | 3.1 | 2.8 |
| $V_{m_L}$ | Maximum vaccinated, low SES | 0.72 | 0.79 | 0.78 |
| $V_{m_H}$ | Maximum vaccinated, high SES | 0.97 | 1 | 0.93 |
| $W_{h_L}$ | Halfway week, low SES | 21.3 | 30.6 | 32.1 |
| $W_{h_H}$ | Halfway week, high SES | 19.9 | 24.5 | 23.4 |

Table S2: Country- and SES-specific vaccination trends.

| Parameter | Symbol | Value | Reference |
|---|---|---|---|
| Probability of infection given contact | | | |
| Wild type | $\beta$ | 18.9% | [7] |
| Delta | $\beta$ | 29.7% | [7] |
| Omicron | $\beta$ | 42.7% | [7] |
| Omicron* | $\beta$ | 46.97, 55.5% | |
| Scale infectiousness for secondary infections | $si$ | 0.27 | [22] |
| Incubation period | $\frac{1}{\epsilon}$ | 4 days | [50] |
| Fraction symptomatic | $\nu$ | 0.4 | [51–53] |
| Infectious Period | $\frac{1}{\gamma}$ | 7 days | [54] |
| Natural immunity[1] | $\frac{1}{\omega}$ | 10 months | [55, 56] |
| Case fatality rate | | | [4, 52] |
| Children | $\rho_{ch}$ | High SES: 0.00044 | |
| | $\rho_{cl}$ | Low SES: 0.001364 | |
| Adults | $\rho_{ah}$ | High SES: 0.00356 | |
| | $\rho_{al}$ | Low SES: 0.00854 | |
| Older adults | $\rho_{eh}$ | High SES: 0.0346 | |
| | $\rho_{el}$ | Low SES: 0.0346 | |
| HSM-specific | | | |
| Scale down protection after waning of immunity | | | |
| 1 immune event | $d_{\omega,1}$ | 0.4 | |
| 2 immune events | $d_{\omega,2}$ | 0.7 | |
| 3+ immune events | $d_{\omega,3}$ | 0.85 | |
| Protection against infection relative to severe disease | $\phi$ | 80% | |
| HIM-specific | | | |
| Scale infection probability to match HSM force of infection | | | |
| Low SES | $\beta_L$ | 9.34% | |
| High SES | $\beta_H$ | 5.34% | |
| Vaccine effectiveness | | | |
| Two doses, no booster | | | [42, 43, 45] |
| Infection effectiveness | $VE_i$ | 0 | |
| Severe disease effectiveness | $VE_h$ | 0.7 | |
| Two doses, with monovalent booster | | | [42–45] |
| Infection effectiveness | $VE_i$ | 0.5256 | |
| Severe disease effectiveness | $VE_h$ | 0.657 | |
| Two doses, with bivalent booster | | | [42–45] |
| Infection effectiveness | $VE_i$ | 0.74 | |
| Severe disease effectiveness | $VE_h$ | 0.925 | |
| Probability of infection given contact ($si$ sensitivity analysis) | | | [23] |
| Low SES | $\beta_L$ | 4.79% | |
| High SES | $\beta_H$ | 1.82% | |

[1] HSM model used same duration of immunity for vaccine and booster-derived immunity.

Table S3: Parameter values used in HSM and HIM simulations.

| Age | Malaysia | Ecuador | India |
|---|---|---|---|
| Child | 21,450 | 23,632 | 24,046 |
| Adult | 66,772 | 63,254 | 64,678 |
| Old | 11,774 | 13,112 | 11,272 |
| Total | 99,996 | 99,998 | 99,996 |

Table S4: HSM simulation sizes by country.

| Contact type | Malaysia | Ecuador | India |
|---|---|---|---|
| Low to Low | 1.53 | 1.55 | 2.89 |
| Low to High | 1.02 | 1.03 | 1.92 |
| High to Low | 0.58 | 0.59 | 1.10 |
| High to High | 0.87 | 0.89 | 1.65 |
| Total contact | 4.00 | 4.06 | 7.56 |

Table S5: HSM baseline contact rates across countries and SES, prior to application of stringency index.

| SAR | country | Scenario | Booster | Relative Benefit |
|---|---|---|---|---|
| 110% | Ecuador | History dependent | Bivalent | 1.18 |
| | | History dependent | Monovalent (Omicron) | 0.84 |
| | India | History dependent | Bivalent | 1.66 |
| | | History dependent | Monovalent (Omicron) | 1.1 |
| | Malaysia | History dependent | Bivalent | 1.12 |
| | | History dependent | Monovalent (Omicron) | 0.89 |
| 130% | Ecuador | History dependent | Bivalent | 1.12 |
| | | History dependent | Monovalent (Omicron) | 0.77 |
| | India | History dependent | Bivalent | 1.77 |
| | | History dependent | Monovalent (Omicron) | 1.04 |
| | Malaysia | History dependent | Bivalent | 1.09 |
| | | History dependent | Monovalent (Omicron) | 0.88 |

Table S6: Relative benefit (Equation 1) of bivalent vs. WT monovalent booster formulation for HSM main text scenarios.

| Country | Analysis | Booster | Relative Benefit |
|---|---|---|---|
| Ecuador | Boost like Malaysia | Bivalent | 1.11 |
| | Boost like Malaysia | Monovalent (Omicron) | 0.82 |
| | Conservative vaccine rollout | Bivalent | 1.25 |
| | Conservative vaccine rollout | Monovalent (Omicron) | 0.76 |
| | Same efficacy scenario | Bivalent | 1.27 |
| | Same efficacy scenario | Monovalent (Omicron) | 1 |
| | Same endpoint scenario | Bivalent | 1.11 |
| | Same endpoint scenario | Monovalent (Omicron) | 1.01 |
| | Vaccinate through Omicron* | Bivalent | 1.19 |
| | Vaccinate through Omicron* | Monovalent (Omicron) | 0.78 |
| India | Boost like Malaysia | Bivalent | 1.42 |
| | Boost like Malaysia | Monovalent (Omicron) | 1.05 |
| | Conservative vaccine rollout | Bivalent | 1.44 |
| | Conservative vaccine rollout | Monovalent (Omicron) | 0.73 |
| | Same efficacy scenario | Bivalent | 1.78 |
| | Same efficacy scenario | Monovalent (Omicron) | 1.19 |
| | Same endpoint scenario | Bivalent | 1.22 |
| | Same endpoint scenario | Monovalent (Omicron) | 0.96 |
| | Vaccinate through Omicron* | Bivalent | 1.68 |
| | Vaccinate through Omicron* | Monovalent (Omicron) | 1 |
| Malaysia | Boost like Malaysia | Bivalent | 1.09 |
| | Boost like Malaysia | Monovalent (Omicron) | 0.88 |
| | Conservative vaccine rollout | Bivalent | 1.12 |
| | Conservative vaccine rollout | Monovalent (Omicron) | 0.84 |
| | Same efficacy scenario | Bivalent | 1.15 |
| | Same efficacy scenario | Monovalent (Omicron) | 1.01 |
| | Same endpoint scenario | Bivalent | 1.07 |
| | Same endpoint scenario | Monovalent (Omicron) | 0.99 |
| | Vaccinate through Omicron* | Bivalent | 1.07 |
| | Vaccinate through Omicron* | Monovalent (Omicron) | 0.86 |

Table S7: Relative benefit (Equation 1) of bivalent vs. WT monovalent booster formulation for supplementary analyses in the HSM. All analyses assume that Omicron* is 30% more infectious than Omicron.

| Parameter | Meaning | Malaysia | Ecuador | India |
|---|---|---|---|---|
| $kB_L$ | Shape of booster trends, low SES | 5.0 | 3.9 | 2.5 |
| $kB_H$ | Shape of booster trends, high SES | 5.5 | 3.1 | 2.8 |
| $B_{m_L}$ | Maximum boosted (excluding children), low SES | 0.91 | 0.87 | 0.35 |
| $B_{m_H}$ | Maximum boosted (excluding children), high SES | 0.90 | 0.88 | 0.35 |
| $WB_{h_L}$ | Halfway week, low SES | 11.3 | 20.6 | 22.1 |
| $WB_{h_H}$ | Halfway week, high SES | 9.9 | 14.5 | 13.4 |

Table S8: Country- and SES-specific booster trends.

| Country | Prior infection | Cross protection |
|---|---|---|
| Ecuador | 78.1% | 0.665 |
| India | 52.4% | 0.433 |
| Malaysia | 90.7% | 0.728 |

Table S9: Country benchmarking parameters based on no booster states estimated from the VSM at the start of the simulation. Cross protection is calculated using the omicron* column of Figure 1A and prior infection is calculated based on final states shown in Figures 1B and S4.

| Parameter | Country | | | Reference |
|---|---|---|---|---|
| | Ecuador | India | Malaysia | |
| Age distribution of infection | | | | [47–49] |
| Child | 53.4% | 35.4% | 36.7% | |
| Adult | 37.8% | 58.7% | 57.8% | |
| Older adults | 8.8% | 6.0% | 5.5% | |
| Population age distribution | | | | |
| Child | 41.0% | 35.5% | 36.2% | |
| Adult | 52.6% | 58.7% | 59.0% | |
| Older adults | 6.4% | 5.8% | 4.8% | |
| Initial reported infections | 1798 | 19153 | 11236 | [46] |
| Reporting rate (symptomatic) | 0.189 | 0.053 | 0.178 | [47–49] |

Table S10: Initial conditions used in Hybrid Immunity Model simulations by country

| Country | SAR | Booster | HIM | HSM | Lower | Upper |
|---|---|---|---|---|---|---|
| Ecuador | 110% | Bivalent | 0.11 | 0.66 | 0.58 | 0.74 |
| | | Monovalent (WT) | 0.07 | 0.56 | 0.48 | 0.65 |
| | | Monovalent (Omicron) | | 0.47 | 0.38 | 0.56 |
| | 130% | Bivalent | 0.38 | 2.88 | 2.69 | 3.08 |
| | | Monovalent (WT) | 0.27 | 2.56 | 2.37 | 2.76 |
| | | Monovalent (Omicron) | | 1.98 | 1.78 | 2.19 |
| India | 110% | Bivalent | 0.57 | 0.68 | 0.56 | 0.81 |
| | | Monovalent (WT) | 0.4 | 0.41 | 0.28 | 0.53 |
| | | Monovalent (Omicron) | | 0.45 | 0.32 | 0.58 |
| | 130% | Bivalent | 0.96 | 2.94 | 2.58 | 3.31 |
| | | Monovalent (WT) | 0.58 | 1.66 | 1.28 | 2.04 |
| | | Monovalent (Omicron) | | 1.72 | 1.34 | 2.11 |
| Malaysia | 110% | Bivalent | 0.11 | 1.35 | 1.26 | 1.43 |
| | | Monovalent (WT) | 0.06 | 1.21 | 1.13 | 1.3 |
| | | Monovalent (Omicron) | | 1.08 | 1 | 1.17 |
| | 130% | Bivalent | 0.22 | 4.48 | 4.32 | 4.63 |
| | | Monovalent (WT) | 0.14 | 4.12 | 3.96 | 4.28 |
| | | Monovalent (Omicron) | | 3.64 | 3.48 | 3.8 |

Table S11: Deaths averted per 10,000 in the HIM versus HSM.
